# Social distancing across vulnerability, race, politics, and employment: How different Americans changed behaviors before and after major COVID-19 policy announcements

**DOI:** 10.1101/2020.06.04.20119131

**Authors:** Vincent Huang, Staci Sutermaster, Yael Caplan, Hannah Kemp, Danielle Schmutz, Sema K. Sgaier

## Abstract

**Background:** As states reopen in May 2020, the United States is still trying to curb the spread of the COVID-19 pandemic. To appropriately design policies and anticipate behavioral change, it is important to understand how different Americans’ social distancing behavior shifts in relation to policy announcements according to individual characteristics, and community vulnerability.

**Methods:** This cross-sectional study used Unacast’s social distancing data from February 24th - May 10th, 2020 to study how social distancing changed before and after: 1) The World Health Organization’s declaration of a global pandemic, 2) White House announcement of “Opening Up America Again” (OUAA) guidelines, and 3) the week of April 27 when several states reopened. To measure intention to social distance, we assessed the difference between weekday and weekend behavior as more individuals have more control over weekend leisure time. To investigate social distancing’s sensitivity to different population characteristics, we compared social distancing time-series data across county vulnerability as measured by the COVID-19 Community Vulnerability Index (CCVI) which defines vulnerability across socioeconomic, household composition, minority status, epidemiological, and healthcare-system related factors. We also compared social distancing across population groupings by race, 2016 presidential election voting choice, and employment sectors.

**Results:** Movement reduced significantly throughout March reaching peak reduction on April 12th (-56.1%) prior the enactment of any reopening policies. Shifts in social distancing began after major announcements but prior to specific applied policies: Following the WHO declaration, national social distancing significantly increased on weekdays and weekends (-18.6% and -41.3% decline in mobility, respectively). Social distancing significantly declined on weekdays and weekends after OUAA guidelines (i.e. before state reopening) (+1.1% and +5.3% increase in mobility, respectively) with additional significant decline after state reopening (+10.0% and +20.9% increase in mobility, respectively). Social distancing was significantly greater on weekends than weekdays throughout March, however, the trend reversed by early May with significantly less social distancing on weekends, suggesting a shift in intent to social distance during leisure time. In general, vulnerable counties social distanced less than non-vulnerable counties, and had a greater difference between weekday and weekend behavior until state reopening. This may be driven by structural barriers that vulnerable communities face, such as higher rates of employment in particular sectors. At all time periods studied, the average black individual in the US social distanced significantly more than the average white individual, and the average 2016 Clinton voter social distanced significantly more than the average 2016 Trump voter. Social distancing behavior differed across industries with three clusters of employment sectors.

**Conclusion:** Both signaling of a policy change and implementation of a policy are important factors that seem to influence social distancing. Behaviors shifted with national announcements prior to mandates, though social distancing further declined nationwide as the first states reopened. The variation in behavioral drivers including vulnerability, race, political affiliation, and employment industry demonstrates the need for targeted policy messaging and interventions tailored to address specific barriers for improved social distancing and mitigation.

## Introduction

According to the Centers for Disease Control and Prevention (CDC), social distancing (maintaining a physical distance of 6 feet between individuals and avoiding group gatherings and crowded spaces) is the best way to help slow the spread of COVID-19 (CDC, 2020). Social distancing is a complex behavior with engagement driven by its relative costs and benefits, both of which may be perceived by individuals as high (Fenichel et al., 2011; Friedson, McNichols, Sabia, & Dave, 2020; Thunström, Newbold, Finnoff, Ashworth, & Shogren, 2020). As COVID-19 cases continue to spread across the United States, social distancing remains an essential mitigation tool. However, many uncertainties remain about how individual social distancing behavior has changed and will change over time, in relation to policy context and individual and community characteristics. To effectively encourage the necessary levels of social distancing for months to come – and to prepare for potential second waves or future pandemics – there is a pressing need to understand the relationship between a person’s social distancing behavior, their characteristics, and what is happening around them.

Research suggests that uptake of social distancing is sensitive to the specific types, timing, and context of policies enacted (Dave, Friedson, Matsuzawa, & Sabia, 2020; Gupta et al., 2020; Lasry A, 2020). To date, state governments have enforced social distancing through a variety of policy actions, such as closing schools and non-essential businesses and placing restrictions on bars, restaurants, and mass gatherings. However, social distancing has been shown to increase not only following the announcement of regulations, as might be expected, but also *prior to* the enactment of policies such as closures of recreation and fitness facilities or the closure of all non-essential businesses (Gupta et al., 2020). Moreover, the level of social distancing varies among states with similar policies, implying that the effectiveness of policies varies with context. Analysis of shelter in place orders, for example, found differences among states in the amount of social distancing that followed the orders, depending on the date the order was enacted and the density of the state’s population (Dave et al., 2020). These heterogeneous findings suggest that the relationship between policy and behavior is complex.

Additionally, to our knowledge, research has not yet explored how people’s social distancing behavior changes after restrictions on movement have been relaxed and states “reopen”. This is critical because the impact of non-social distancing behavior may be masked by the delays between transmission and confirmation of COVID-19 infection (Courtemanche, Garuccio, Le, Pinkston, & Yelowitz, 2020; Davies, Kucharski, Eggo, Gimma, & Edmunds, 2020; Ferguson et al., 2020; Flaxman et al., 2020; Pei, Kandula, & Shaman, 2020). Policymakers need to understand social distancing behavior and intent in order to guide an early targeted response (e.g. re-introduction of social distancing policies, mandated outdoor masks, etc.) to prevent unexpected growth in cases and deaths.

Social distancing behavior may also shift within the course of a week due to drivers unrelated to policy. These within-week differences in behaviors provide insight into how people behave when they have more control over their time (such as on the weekends), a good proxy of social distancing intent. For example, subway use in South Korea was greater on weekdays, when it was needed for essential work-related travel, and less on weekends, suggesting increased social distancing behavior during leisure time (Park, 2020). In the US, the average radius of weekend travel initially remained greater than weekday travel (i.e. people traveled greater distances), but the difference disappeared by mid-March, suggesting an increase in those staying home (Klein et al., 2020). These in-week variations suggest that social distancing behavior on weekdays and weekends should be analyzed separately (rather than looking at a week overall) to better understand shifts in behavior over time and as an indicator of social distancing intent. To date, however, the relationship between weekdays and weekends has not been explored in detail.

Individual and community characteristics influence social distancing uptake as well. Social distancing behavior can be influenced by monetary loss (Bodas & Peleg, 2020), political affiliation of individuals and policymakers (Adolph, Amano, Bang-Jensen, Fullman, & Wilkerson, 2020; Allcott et al., 2020; Andersen, 2020; Painter & Qiu, 2020), and poverty (Wright, Sonin, Driscoll, & Wilson, 2020). Research has shown that different population segments vary in their beliefs and behaviors around social distancing: the segment practicing the least social distancing has low risk perception and low self-efficacy, and is more likely to be male and Republican (Charles et al., 2020). Employees who work in industries that require face-to-face communication or physical proximity with other workers may also be affected differently by social distancing policies (Koren & Pető, 2020).

In addition to these individual characteristics, research has shown that certain geographic communities are better equipped to respond to disasters than others, which suggests that there may also be differences in social distancing uptake based on community vulnerability (Flanagan, Gregory, Hallisey, Heitgerd, & Lewis, 2011). Vulnerable communities may face disproportionate socioeconomic costs, such as job loss and income reductions, as a result of social distancing that make the practice less accessible or more burdensome (Fairchild, Gostin, & Bayer, 2020; Flanagan et al., 2011; Hutchins, Fiscella, Levine, Ompad, & McDonald, 2009; Lewnard & Lo, 2020). While existing research identifies several individual factors that influence social distancing behavior, there is no comprehensive assessment that characterizes social distancing across these factors.

As states continue to reopen across the nation, it is essential for national and state policymakers to understand local social distancing behavior to guide their responses to the continued spread of the virus. Examining this heterogeneity in how communities respond to shifting policies and how these responses vary by community and individual characteristics may help policymakers better balance trade-offs, such as between the economic benefits of reopening and the continued need for social distancing, as well as determine what messaging to use in their policy announcements.

This case study leverages large-scale, near real-time mobility data to explore national social distancing trends over time, as well as how these changes differ across population subgroups. Our analysis uses a COVID-19 Community Vulnerability Index (CCVI) to explore the relationships between social distancing and the factors that make a community less able to cope with the impact of the pandemic (Surgo Foundation, 2020a). We address four questions: First, how much are Americans social distancing and how has this changed over time? Second, how has mobility increased or decreased in the weeks following key policy announcements? Third, as most business-related activities are conducted during weekdays, what is the difference in mobility levels between weekends versus weekdays and how has this difference changed over time? Fourth, how do these temporal trends in social distancing differ across population subgroups, including vulnerability, political affiliation, race, and employment sector? Results have important implications for policymakers’ continued adaptation of social distancing and other measures to fight COVID-19.

## Methods

### Data Sources

#### Social Distancing Data

Near real-time social distancing data were gathered at the county level from Unacast’s analysis of mobile phone location data (Unacast, 2020). Data were collected from over 2,500 mobile phone applications, from February 24th to May 10^th^, 2020, inclusive. Social distancing is reflected as a percentage change in the distance travelled when compared to pre-pandemic averages (for any given day of the week, the pre-pandemic average is defined by Unacast as the average distance traveled on the four same days of the week between February 10th and March 8th 2020, inclusive). A negative number represents more social distancing, and a positive number less social distancing. For example, if individuals in a county average 10 miles of travel on the four Wednesdays in pre-pandemic days, but only average 8 miles on a Wednesday during the pandemic, a -20% measurement is recorded as the mobility decline, i.e., degree of social distancing, for the county on that date.

#### National Announcements and State Reopening Policies

National policy announcements were selected based on their prevalence in news coverage of national COVID-19 timelines. Announcements that featured in multiple timelines were selected as indicating major shifts in the pandemic discourse. Effective dates of state reopening policies were tracked over time from the Kaiser Family Foundation and National Governors Association resources and confirmed with local government documentation of signed executive orders (Kaiser Family Foundation, 2020; National Governors Association, 2020).

#### County Characteristics

To understand how social distancing might be sensitive to vulnerability within the context of the coronavirus pandemic, we used the COVID-19 Community Vulnerability Index (CCVI, to be published) to identify communities that have a limited ability to delay and mitigate the health, economic, and social impacts of a pandemic. Recognized by the CDC, the index builds on the CDC’s own Social Vulnerability Index (SVI) (Flanagan et al., 2011). The CCVI’s 34 indicators are grouped into 6 core themes that reflect a community’s vulnerability to the COVID-19 pandemic: the four existing themes from the SVI including socioeconomic status, household composition and disability, minority status and language, and housing type and transportation; and two themes specific to COVID-19 - epidemiological factors, and healthcare-system factors. Data are available online at <u>The COVID-19 Community Vulnerability Index (CCVI)</u> along with a detailed methodology <u>(CCVI Methodology</u>) (Surgo Foundation, 2020a, 2020b). The CCVI and its component thematic scores are on a percentile scale, and we grouped counties into least vulnerable (x < 0.33), moderately vulnerable (0.33 <= x < 0.67), or most vulnerable (x >= 0.67), where x is either the county’s aggregate CCVI score or any of its component thematic scores.

Data on race were sourced from the US Census Bureau’s 2018 American Community Survey (US Census Bureau, 2018). For each county, the Census Bureau estimated the number of residents who identify with one or more race categories, including American Indian or Alaska Native, Asian, Black, Native Hawaiian or Other Pacific Islander, or White. Hispanic or Latino origin was not included because the Census Bureau classifies this separately as ethnicity rather than race. For simplicity, only census categories “Black alone” (labeled “black” in our analysis) and “White alone” (labeled “white”) were analyzed; other racial groups were excluded because of their low prevalence in the population relative to other races.

To measure political affiliation, voting data from the 2016 Presidential election were sourced from the MIT Election Data and Science Lab (MIT Election Data and Science Lab, 2018). The data set includes vote counts for the Republican nominee, Donald Trump, and for the Democratic nominee, Hillary Clinton, for each county in the US. The exception is Alaska, which reported counts by districts at the time of the election and not by county-equivalent borough boundaries. Boroughs without corresponding district reports were excluded from the MIT data set and the current analysis.

Employment-sector data were sourced from the US Bureau of Labor Statistics’ Quarterly Census of Employment and Wages, which tracks employment numbers per job sector (US Bureau of Labor Statistics, 2019). To account for seasonality, we used March 2019 data to approximate employment in each sector in the months prior to the pandemic. The total number of employees employed by private establishments was aggregated into 20 employment sectors (2-digit sector code level, as defined by the North American Industry Classification System) per county. To simplify our analysis, we did not include government employees, which also led to the exclusion of the public administration sector.

#### COVID-19 Case Data

Confirmed cases and death counts were sourced from the Johns Hopkins University Center for Systems Science and Engineering, which collects and reports COVID-19 data for each US county (Dong, Du, & Gardner, 2020). Because counties occasionally switch reporting formats, reported cumulative counts may decrease at times. We rectify this by enforcing the cumulative counts to be monotonically increasing to only report peak cumulative counts – if the cumulative count is less than the previous day, the data is replaced with the previous day’s count.

#### Data availability, consent, and ethics statement

All data are publicly available, other than social distancing data obtained through Unacast’s Data for Good initiative. All data sets are deidentified. We received Unacast’s deidentified social distancing data already aggregated at the level of counties, which cannot be traced to individuals. Unacast has an explicit privacy and consent policy (https://www.unacast.com/opt-out) stating that mobile phone users have opt-in consent for the collection of location data from mobile devices. Since the current study is a secondary analysis of existing, deidentified datasets obtained at the county-level, it did not require IRB approval. Surgo Foundation approved the study.

### Analysis

We analyzed national social distancing data as time series for time periods before and after specific events, weekdays and weekends, and population groupings. For analysis without population groupings, social distancing was aggregated at the national level by computing the weighted mean from the county social distancing data, based on county population. The full analyses were conducted for all US counties at the national level. The dates of key national announcements were graphed with social distancing along with COVID cases and deaths per population from February 24th to May 10th, 2020 (last day of social distancing data availability).

#### Comparisons across population groupings

We characterized how much social distancing was achieved by different population groupings across community vulnerability, race, 2016 presidential election voters, and employment sector. Characteristics were selected based on a review of existing evidence on potential influences on social distancing compliance (Adolph et al., 2020; Allcott et al., 2020; Andersen, 2020; Bodas & Peleg, 2020; Charles et al., 2020; Fairchild et al., 2020; Flanagan et al., 2011; Hutchins et al., 2009; Koren & Pető, 2020; Lewnard & Lo, 2020; Painter & Qiu, 2020; Wright et al., 2020). The groupings were made along the following dimensions, and mean social distancing was computed as described below:

1. **Overall vulnerability to cope with COVID-19:** We assessed social distancing of the most vulnerable, moderately vulnerable, and least vulnerable counties according to the CCVI to compare social distancing by vulnerability level. There is one CCVI score per county, so we computed the mean at the national level by weighting each county’s social distancing by its respective population.
2. **Thematic vulnerability to COVID-10:** The CCVI divides overall vulnerability into six component themes: **1) socioeconomic factors, 2) household composition and disability, 3) minority status and language, 4) housing and transportation, 5) epidemiological factors, and 6) healthcare-system factors**. For each of these component themes, we assessed social distancing of the most vulnerable, moderately vulnerable, and least vulnerable counties, aggregated at the national level. There is one score per thematic vulnerability per county, so we computed the mean at the national level by weighting each county’s social distancing by its respective population.
3. **Community racial make-up:** We estimated how much social distancing was done by black and white Americans. The Census Bureau provides the estimated sub-population by race in a county. We computed the mean social distancing by race at the national level by weighting each county’s social distancing by its respective population estimates per race.
4. **Political affiliation:** We estimated how much social distancing was conducted by populations that voted for Donald Trump or Hillary Clinton in the 2016 presidential election. We repeated the same process used for the racial analysis to compute the weighted mean social distancing per voting group.
5. **Employment sectors:** We estimated how much social distancing was observed in aggregate by 20 employment sectors. We did this by computing social distancing for individual employment sectors at the national level, by weighting county social distancing by each sector’s employment level (i.e., number of jobs) in the county.

#### Comparisons over time

For each population grouping listed above, we analyzed three key events: (1) the World Health Organization (WHO) declaration of a global pandemic on March 11th, 2020, (2) the release of President Trump’s national guidelines for reopening (“Opening Up America Again” – abbreviated OUAA) on April 16th, and (3) the time period (including effective date) of states’ first relaxation of social distancing policies. For each event, we selected a “before” and “after” period for comparison, defined as the first full week before and the first full week after the event of interest. For events 1 and 2, these are the immediate full weeks (Monday-Sunday) before and after the event, without overlapping with the week of the event. For event 3, because the first states to reopen did so during the weeks overlapping with the OUAA release period, to avoid overlap in comparison periods, we compared the week following OUAA with the last available full week of mobility data (the week of May 10th at the time of analysis). During this time period, 16 states relaxed restrictions (and an additional 10 did so in the weeks prior), making it a reasonable proxy for comparing movements before and after state reopenings (Appendix Table 1). Table 1 lists the dates for the periods of interest.

#### Comparisons between weekdays and weekends

We evaluated weekday and weekend trends separately because social distancing graphs show clear differences in weekday and weekend movement, likely because most business activities are conducted during weekdays, even after closure announcements. The percentage change in movement (i.e. social distancing) was averaged for weekdays (Monday to Friday) and for weekends (Saturday and Sunday). The percentage change in social distancing during each time period (Table 1) was calculated by comparing the average social distancing on weekdays before and after each event, and similarly for weekends. In addition, for each week used for the before-and-after event comparisons, the difference in weekday and weekend social distancing was assessed by calculating the percentage change in social distancing on the average weekday (Monday to Friday) compared with weekend (Saturday and Sunday) for the time periods listed in Table 1. For categorical analyses, for both 1) comparisons between periods before and after key events, and 2) differences between weekdays and weekends, the magnitude of the percentage change in social distancing was compared across the five dimensions outlined.

**Table 1.** Event Comparison Periods.

| Time Period | Weekday Comparison |  | Weekend Comparison |  |
| --- | --- | --- | --- | --- |
|  | Before | After | Before | After |
| March 11th WHO Pandemic Announcement | March 2nd - 6th | March 16th-20th | March 7-8th | March 21-22nd |
| April 16th announcement of President Trump’s national guidelines for reopening | April 6th-10th | April 20th-24th | April 11th-12th | April 25th-26th |
| State-level relaxation policies | April 20th-24th | May 4th-8th | April 25th-26th | May 9th-10th |

Since interaction effects are expected between population groupings, for each event a repeated measure, type III Analysis of Variance (ANOVA), was conducted to assess the interaction effects on social distancing of (1) before vs after key events, (2) weekend vs weekdays, and (3) categories within the population grouping. Where interaction effects are identified with an alpha value of 0.05, we conducted post-hoc analyses using paired, weighted, two-tailed Student t-tests, to determine whether there was a significant change in the amount of social distancing before and after the event in interest, between weekdays and weekends in the same seven-day period, or between population grouping categories within each time period.

#### Clustering of employment sectors based on social distancing similarity

We examined social distancing as observed for each employment sector. To further classify variations in social distancing among different sectors, we used k-mean clustering, an unsupervised classification algorithm, to group employment sectors into three clusters on a given day based on how close (or far away) the sectors’ social distance measures were to each other on that day. We then tallied how many times a sector was classified in each cluster and assigned the sector to the cluster with the highest frequency.

## Results

### Social distancing peaked on and declined after April 12th, prior to the enactment of any reopening policies

Nationally there was an overall decrease in mobility (i.e. an increase in social distancing) corresponding with the beginning of the COVID-19 pandemic in the US in early March (Figure 1). Throughout March, mobility declined, indicating that social distancing was increasing with the number of confirmed cases. However, the magnitude of the decline in mobility peaked nationally on April 12th, with 56.1% less mobility recorded than prior to the pandemic. Following this peak, social distancing decreased, despite a continued increase in new cases.

**Figure 1.**
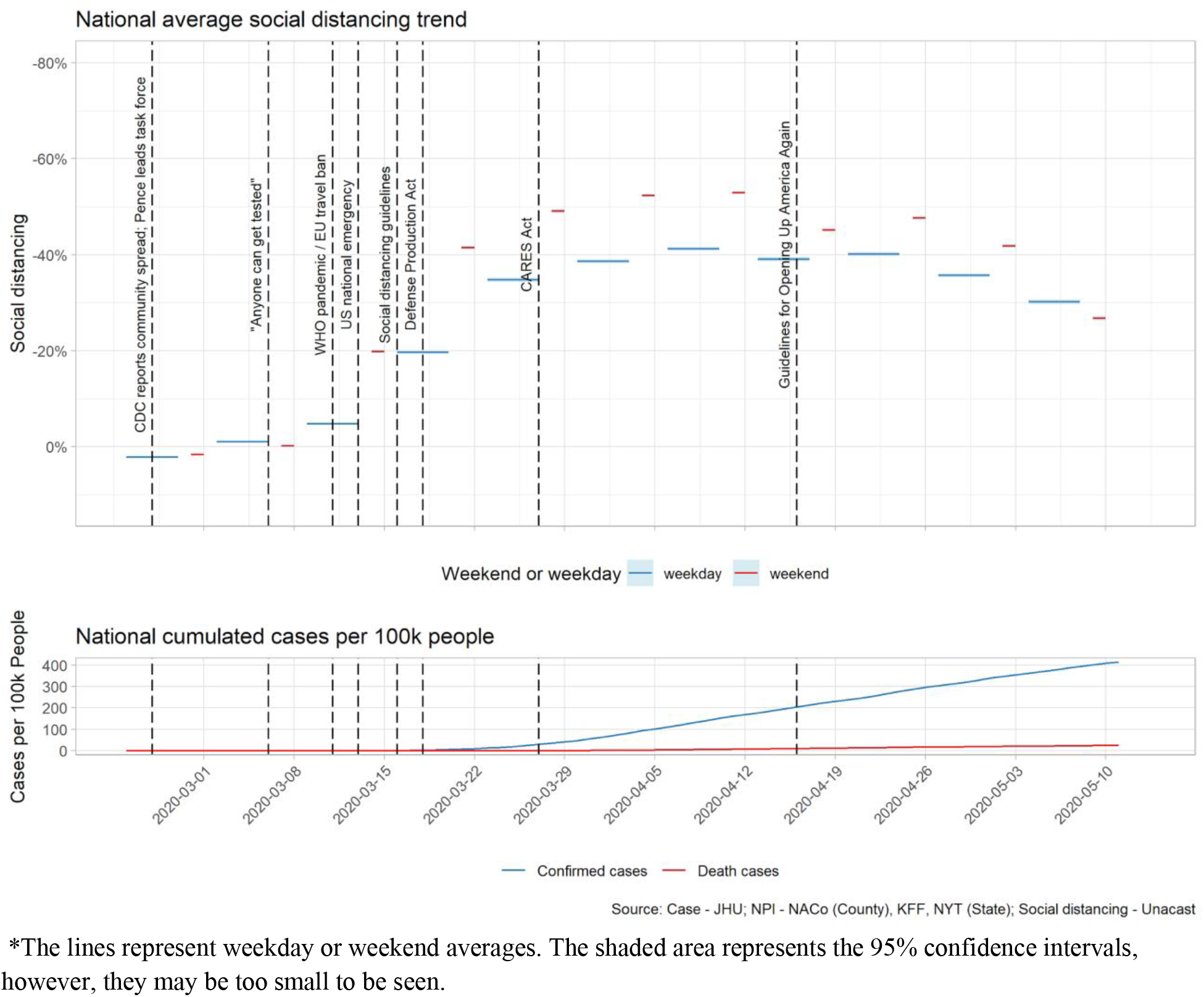
National Average Social Distancing and Cumulative COVID-19 Cases and Deaths.

### Shifts in social distancing began prior to specific applied policies and after major announcements

During the week of March 16th, following the WHO declaration of a COVID-19 pandemic on March 11th and President Trump’s declaration of a national emergency on March 13th, national social distancing significantly increased both on weekdays – with a 18.6% decline in mobility (p>0.01) compared with the week of March 2nd – and weekends – with a 41.3% decline (p>0.01) (Table 2). This increase in social distancing occurred before the CDC announced specific social distancing guidelines on March 16th. In the week beginning April 20th, after the White House had released the OUAA guidelines, individuals socially distanced significantly less on weekdays (1.1%, p>0.01 less social distancing) and on the weekends (5.3%, p>0.01 less) than during the week prior to the week of the guideline release. This decline (i.e., increase in mobility) occurred before any states officially relaxed social distancing policies, which were not implemented until the week of April 27th. Following the first state reopenings, during the week of May 4th, national social distancing significantly declined further, with 10.0% (p>0.01) less social distancing on weekdays and 20.9% (p>0.01) less on weekends, compared with the week prior to relaxed social distancing mandates. This trend was observed regardless of reopening date (Appendix Figure 1).

**Table 2.** National Changes in Mobility (Social Distancing) across Three Time Periods for Average Weekdays and Weekends. *Negative numbers represent a decrease in mobility, i.e. more social distancing than before; positive numbers represent an increase in mobility, i.e. less social distancing than before*.

| WHO Pandemic Declaration |  | OUAA Guidelines |  | State Reopening |  |
| --- | --- | --- | --- | --- | --- |
| Weekday | Weekend | Weekday | Weekend | Weekday | Weekend |
| %Δ | %Δ | %Δ | %Δ | %Δ | %Δ |
| -18.6* | -41.3* | 1.1* | 5.3* | 10.0* | 20.9* |

\*Significant at $p<0.05$ ; otherwise <sup>(ns)</sup>, not significant

### Social distancing was significantly greater on weekends than weekdays after the pandemic declaration, but the gap between the two lessened and disappeared by late April

In the week of March 2nd, prior to the March 11th WHO pandemic announcement, there was a significant small-magnitude difference between weekday and weekend social distancing: national social distancing was 0.9% (p>0.01) greater on weekdays than on weekends (Table 3). By the week of March 16th, following the pandemic announcement, 21.8% (p>0.01) more social distancing occurred on weekends compared with weekdays. Throughout April, social distancing remained higher on weekends than weekdays, although the magnitude of the disparity declined from early to late April, being 11.7% and 7.5% for the weeks of April 6th and 20th, respectively. However, by the week of May 4th, the first week following state reopening, the trend reversed: national social distancing was now 3.4% (p>0.01) greater on weekdays than weekends.

**Table 3.** Difference in Level of Social Distancing between Average Weekend and Weekdays across Five Time Periods. *Negative numbers represent less mobility, i.e., more social distancing, on weekends than weekdays; positive numbers represent more mobility, i.e., less social distancing, on weekends than weekdays*.

| WEEK 1<br>Before WHO<br>Pandemic<br>Announcement | WEEK 2<br>After WHO<br>Pandemic<br>Announcement | WEEK 3<br>Before OUAA<br>Guidelines | WEEK 4<br>After OUAA<br>Guidelines | WEEK 5<br>After Reopening |
| --- | --- | --- | --- | --- |
| %Δ | %Δ | %Δ | %Δ | %Δ |
| 0.9* | -21.8* | -11.7* | -7.5* | 3.4* |

\*Significant at $p<0.05$ ; otherwise <sup>(ns)</sup>, not significant

### More vulnerable counties consistently social distance less

Vulnerable counties practiced significantly less social distancing than non-vulnerable counties at all time periods studied (Figure 2). On April 12th, all three vulnerability groups reached peak social distancing; the decrease in mobility was greatest for least-vulnerable counties (58.4%, 95% CI [57.8%, 59.0%]), significantly less so for moderately vulnerable counties (55.5%, 95% CI [55.0%, 56.1%]), and even less so in the most-vulnerable counties (55.0%, 95% CI [54.3%, 55.8%]). The difference in peak social distancing between moderately vulnerable and most-vulnerable counties is not significant. Nationally, all three vulnerability groups followed the same trends in making significant changes in social distancing after key announcements and state reopenings (Table 4). However, the magnitude of change differed by vulnerability. The interaction effects of vulnerability on the change in social distancing before and after all three key events were significant (F(2,3046)=87.8, p<0.01), (F(2,3046)=38.0, p<0.01), (F(2,3046)=34.4, p<0.01). While most-vulnerable counties increased social distancing on weekdays and weekends following the WHO pandemic declaration, with decreases in mobility of 14.4% and 36.9% (p<0.01), respectively, they social distanced less than least-vulnerable counties, where mobility decreased by 21.8% and 42.7% (p<0.01), respectively. Likewise, although social distancing declined generally following the OUAA announcement on March 16th, least-vulnerable counties saw less of a decline on both weekdays and weekends (mobility increased by 0.6% and 4.5%, p<0.01 respectively) than most-vulnerable counties (mobility increased by 1.8% and 7.1%, p<0.01 respectively). Following state reopenings, most-vulnerable counties experienced a greater decline in social distancing on weekdays and weekends (11% and 20.8% more mobility, p<0.01) compared with least-vulnerable counties (9.1% and 20.2%, p<0.01).

National trends in the differences between weekday and weekend social distancing were similar for all vulnerability groups with significant interaction effects for all time periods (F(2,3046)=36.8, p<0.01), (F(2,3046)=67.9, p<0.01), (F(2,3046)=65.7, p<0.01). However, most-vulnerable counties had a greater disparity between weekday and weekend behavior. This trend disappeared in the week after reopenings, with most-vulnerable counties having a smaller magnitude in weekday and weekend differences compared with moderately and least-vulnerable counties (Table 5).

**Figure 2.**
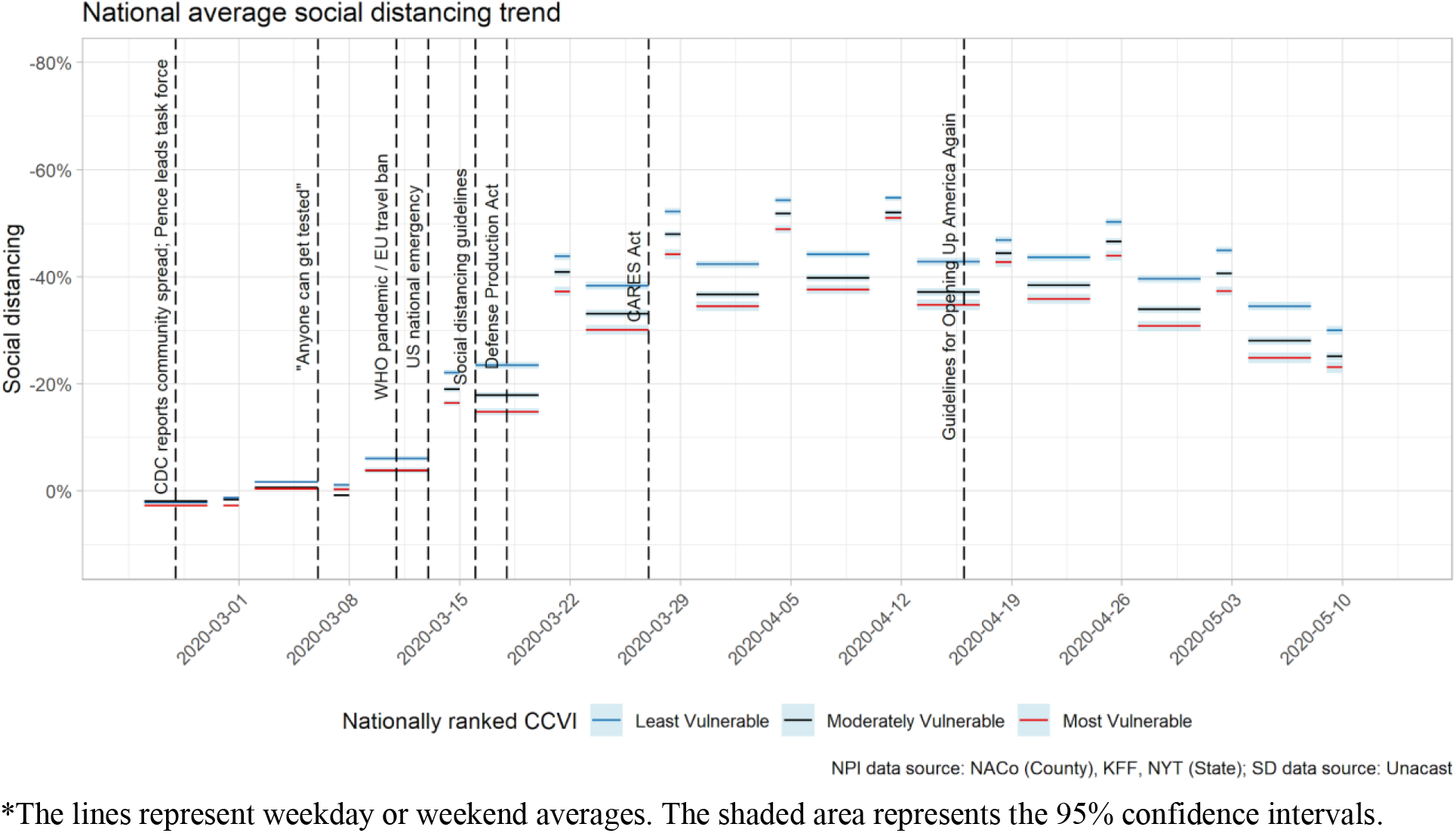
National average social distancing by CCVI vulnerability score.

**Table 4.**
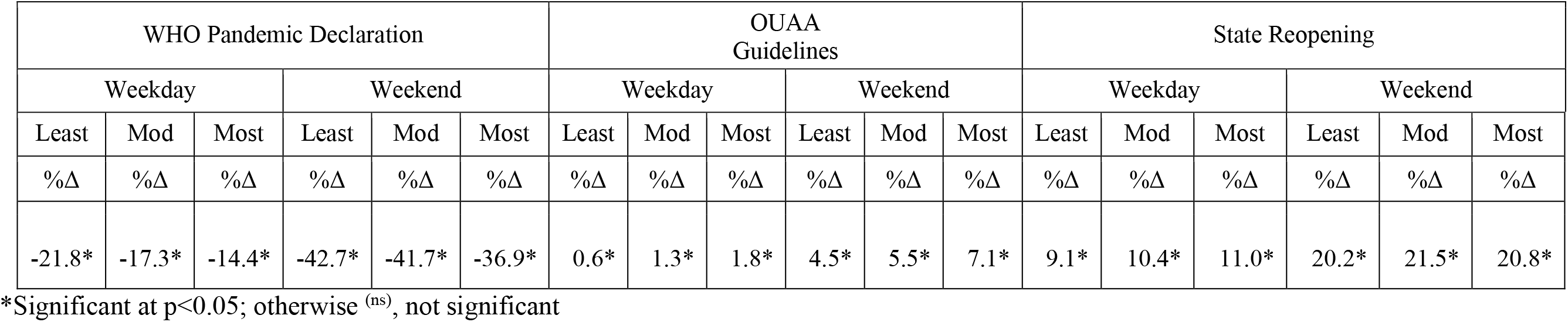
National Changes in Mobility (Social Distancing) across Three Time Periods for Average Weekdays and Weekends, by CCVI Score. *Negative numbers represent a decrease in mobility, i.e. more social distancing than before; positive numbers represent an increase in mobility, i.e. less social distancing than before*.

**Table 5.** Difference in Level of Social Distancing between Average Weekend and Weekdays across Five Time Periods, by CCVI Score. *Negative numbers represent less mobility, i.e., more social distancing, on weekends than weekdays; positive numbers represent more mobility, i.e., less social distancing, on weekends than weekdays*.

| WEEK 1<br>Before WHO Pandemic<br>Announcement |  |  | WEEK 2<br>After WHO Pandemic<br>Announcement |  |  | WEEK 3<br>Before OUAA Guidelines |  |  | WEEK 4<br>After OUAA Guidelines |  |  | WEEK 5<br>After Reopening |  |  |
| --- | --- | --- | --- | --- | --- | --- | --- | --- | --- | --- | --- | --- | --- | --- |
| Least | Mod | Most | Least | Mod | Most | Least | Mod | Most | Least | Mod | Most | Least | Mod | Most |
| %Δ | %Δ | %Δ | %Δ | %Δ | %Δ | %Δ | %Δ | %Δ | %Δ | %Δ | %Δ | %Δ | %Δ | %Δ |
| 0.6* | 1.4* | 0.1<sup>(ns)</sup> | -20.3* | -22.9* | -22.5* | -10.5* | -12.2* | -13.4* | -6.6* | -8.1* | -8.1* | 4.5* | 3.0* | 1.8* |

\*Significant at $p < 0.05$ ; otherwise <sup>(ns)</sup>, not significant

### The level of social distancing differed by types of vulnerability across socioeconomic, demographic, and household composition factors but the difference was more nuanced on epidemiological, health system factors

The relationship between the six themes that compose the CCVI and social distancing varied by vulnerability type. Socioeconomic vulnerability (Figure 3a) and household composition vulnerability (Figure 3b) followed the same pattern as overall vulnerability: counties that were most vulnerable based on these themes social-distanced significantly less than less-vulnerable counties at all time points studied. The same was true for housing and transport vulnerability (Figure 2c) and healthcare-system vulnerability (with the exception of the weekends before and after the WHO announcement) (Figure 2d), but the magnitude of differences between vulnerability categories are smaller. For epidemiological vulnerability (Figure 3e), the difference in social distancing was only significant during the time period around the WHO announcement (F(2,3047)=11.7, p<0.01) and the weekdays (F(2,3047)=.57, p=0.55), but not for the weekends before and after the OUAA guidelines release. By the weeks before and after state reopenings, there was no significant difference in social distancing by epidemiological vulnerability level (Figure 3e). For minority status and language vulnerability the pattern was generally reversed: counties that are more vulnerable based on this theme social distanced more (Figure 3f). This interaction effect was significant at all time points (F(2,3047)=179,p<0.01), (F(2,3047)=107.6, p<0.01), (F(2,3047)=13.87, p<0.01).

Socioeconomic vulnerability, household composition vulnerability, housing and transport vulnerability, and health-system factor vulnerability (with the exception of weekends after state reopenings) all follow the same overall vulnerability trends in the differences in magnitude, significance, and direction of change in social distancing during each time period (Table 6). For both weekdays and weekends, most-vulnerable counties in these themes increased social distancing less than less-vulnerable counties following the WHO announcement, and reduced social distancing more than them following the OUAA guidelines and state reopenings. Epidemiologically vulnerable counties increased their social distancing less than counties with low epidemiological vulnerability following the WHO pandemic announcement, but there was no significant difference in the changes in social distancing by vulnerability for the other time periods for this theme. Counties vulnerable by minority status and language followed an opposite trend: most-vulnerable counties in this theme increased social distancing more after the WHO pandemic announcement than less-vulnerable counties, and decreased social distancing less following the OUAA guidelines and state reopenings. The only exception was on weekends after state reopenings, where most-vulnerable counties in this theme decreased social distancing more than less-vulnerable counties. There is heterogeneity by theme for within week differences between weekday and weekend social distancing behavior over time (Table 7).

**Figure 3a.**
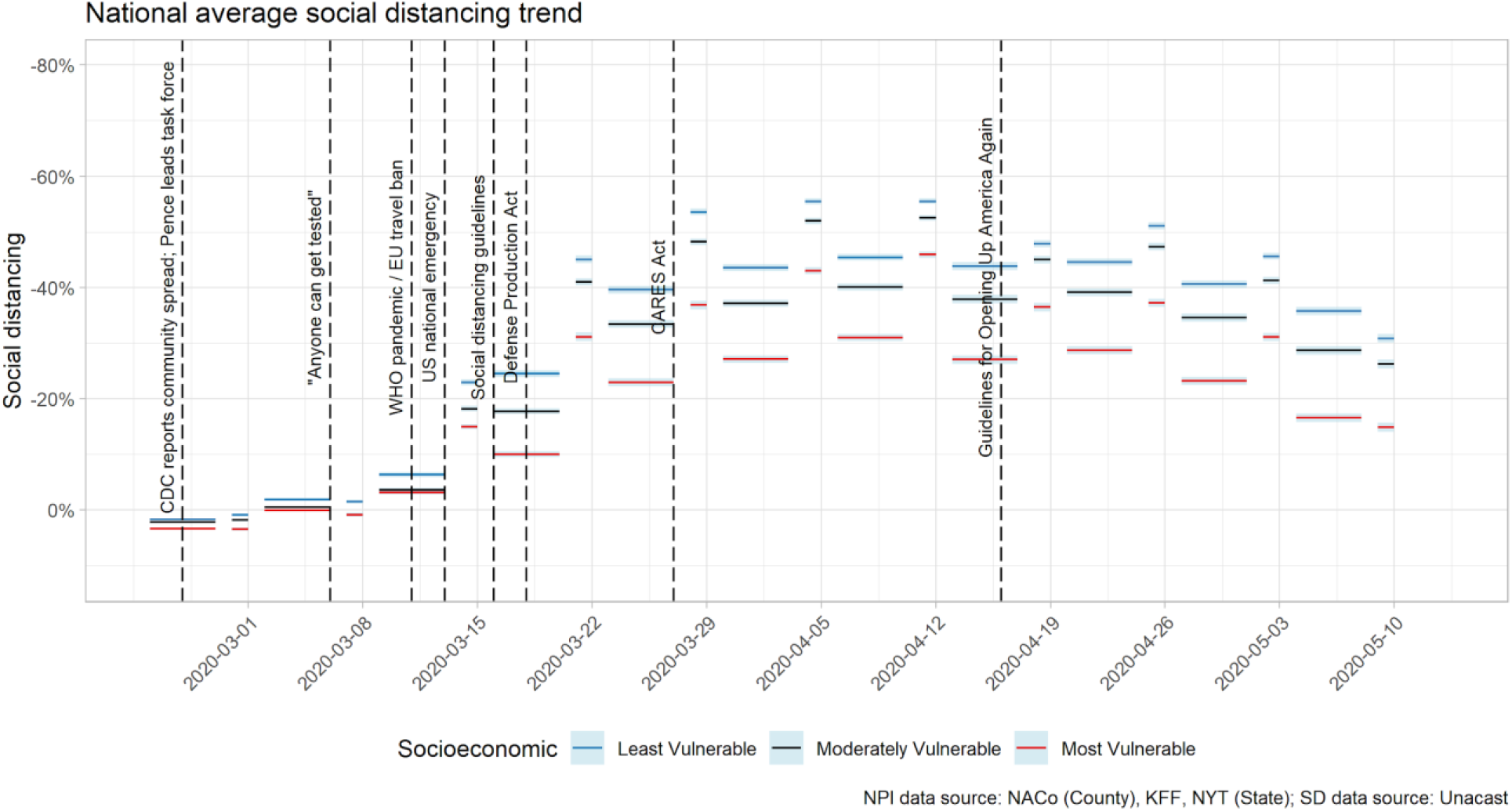
National average social distancing by CCVI socioeconomic status score.

**Figure 3b.**
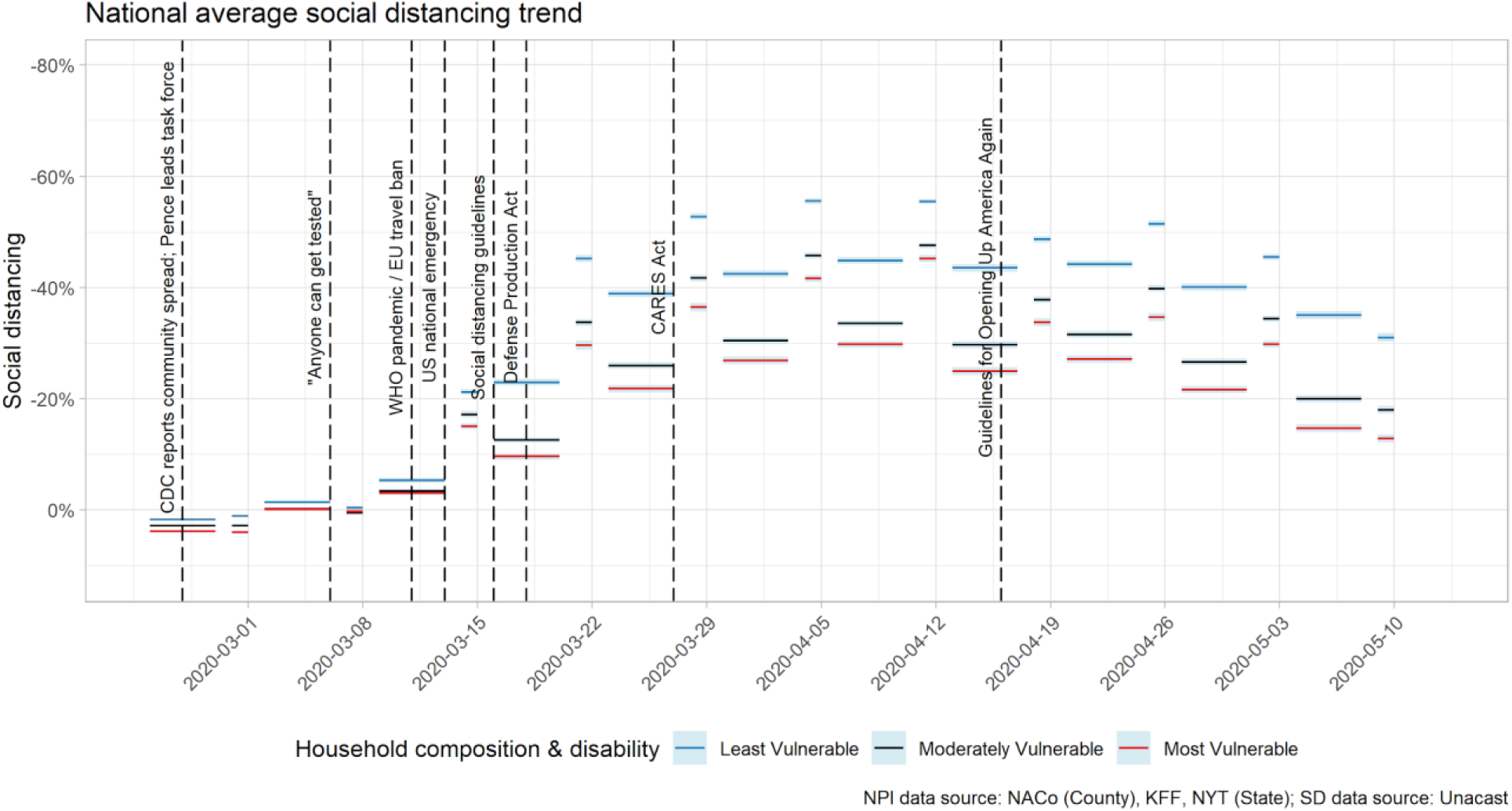
National average social distancing by CCVI household composition & disability score.

**Figure 3c.**
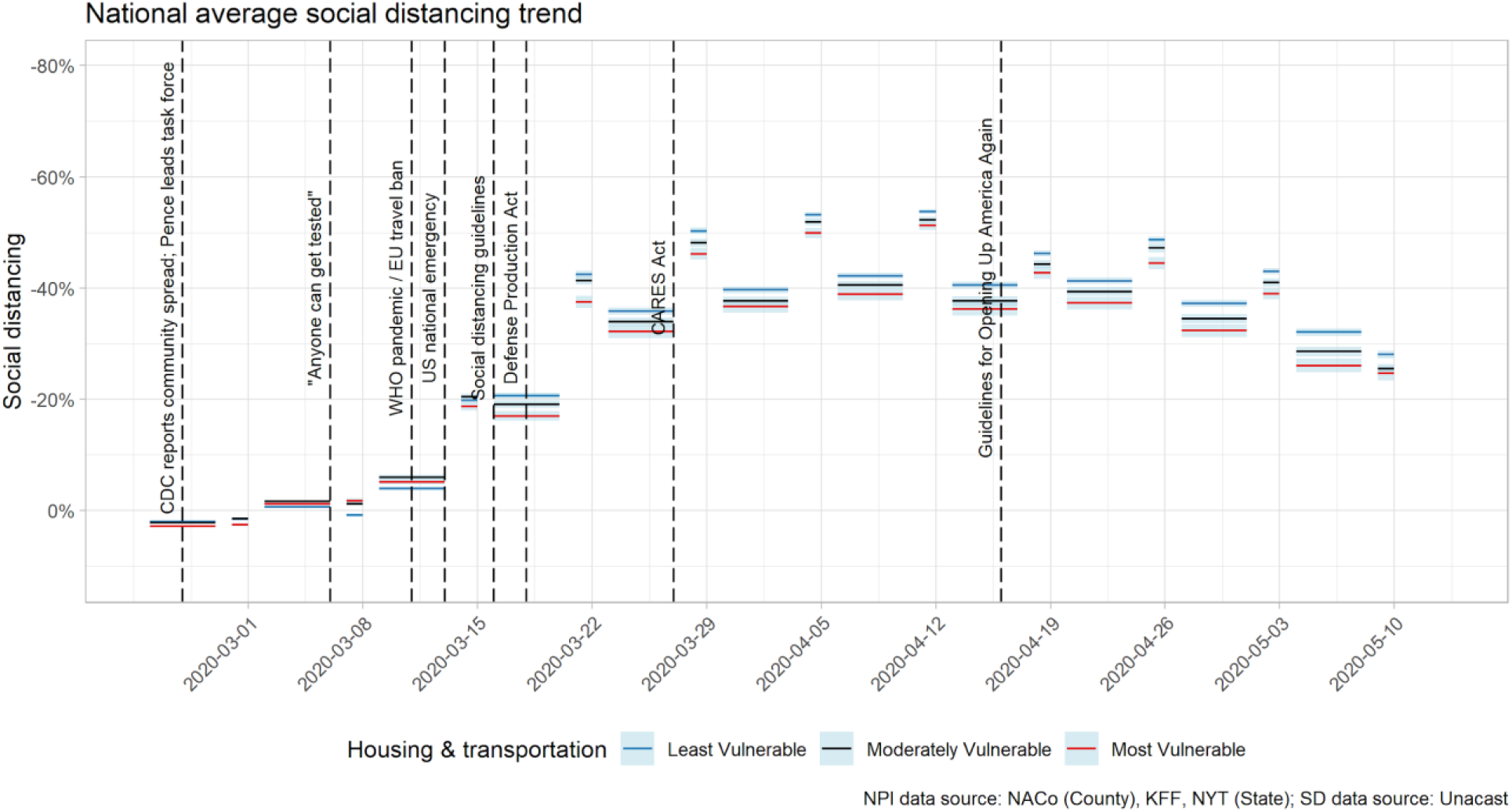
National average social distancing by CCVI housing & transportation score.

**Figure 3d.**
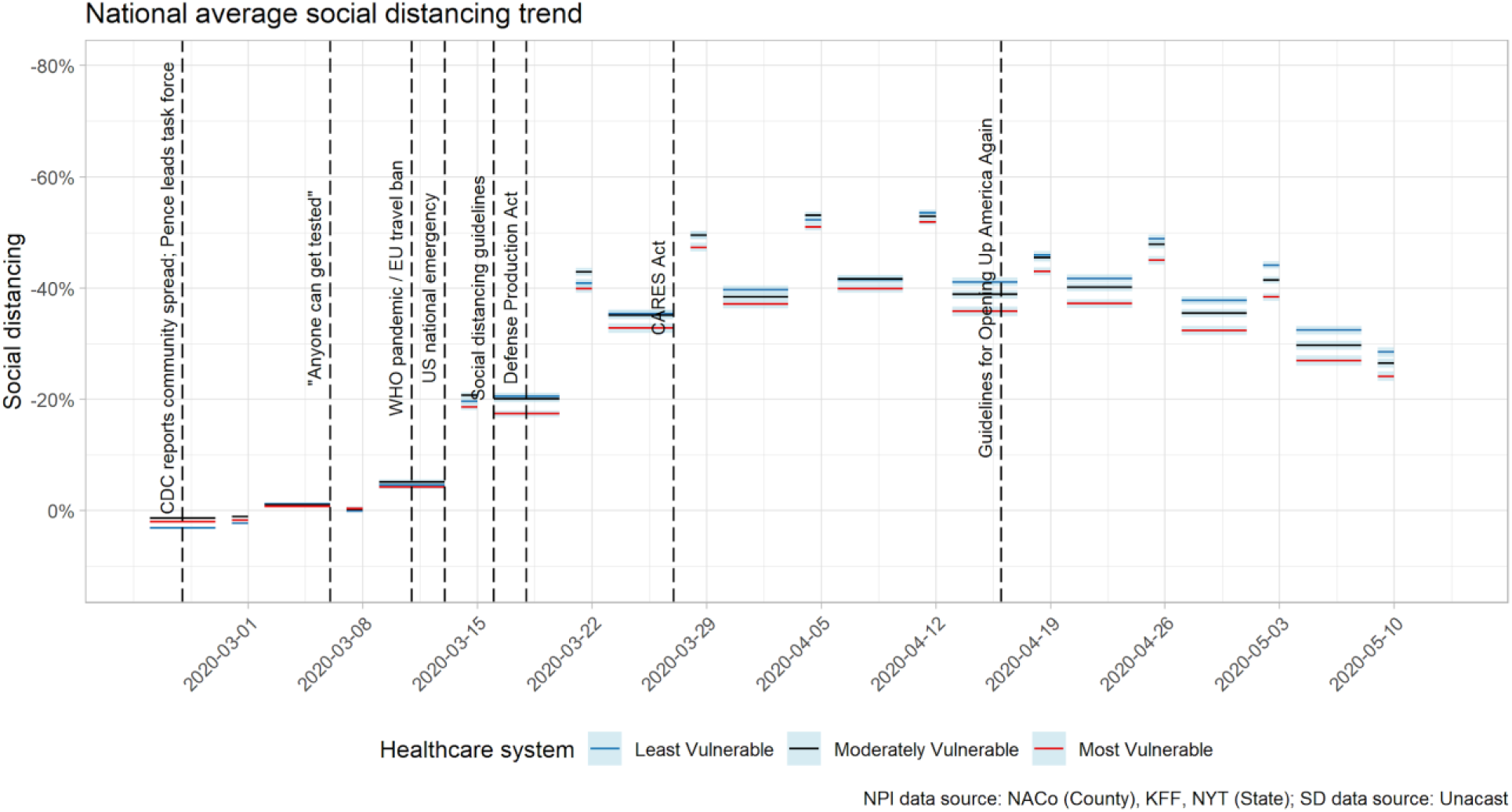
National average social distancing by CCVI healthcare system factors score.

**Figure 3e.**
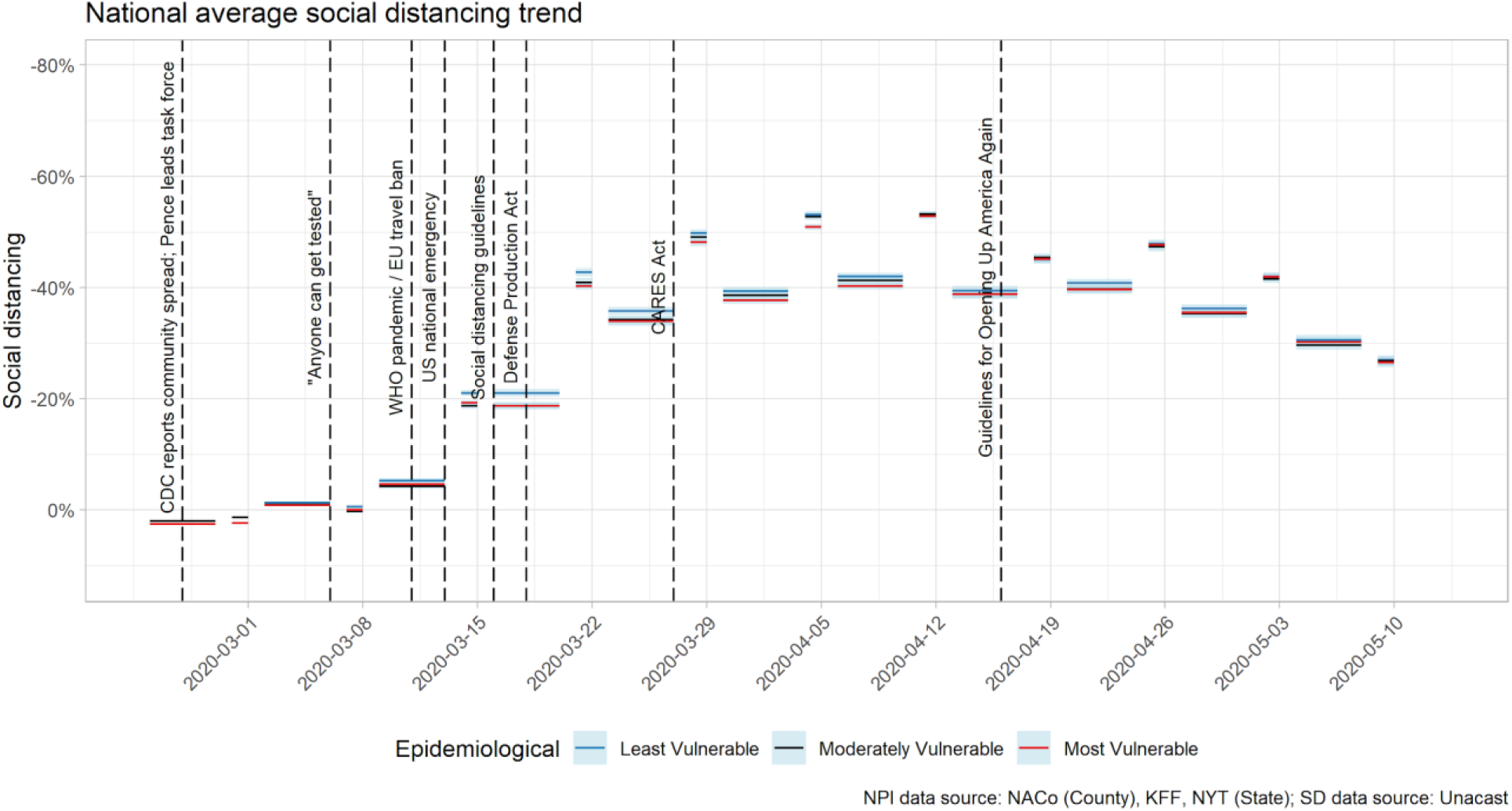
National average social distancing by CCVI epidemiological factors score.

**Figure 3f.**
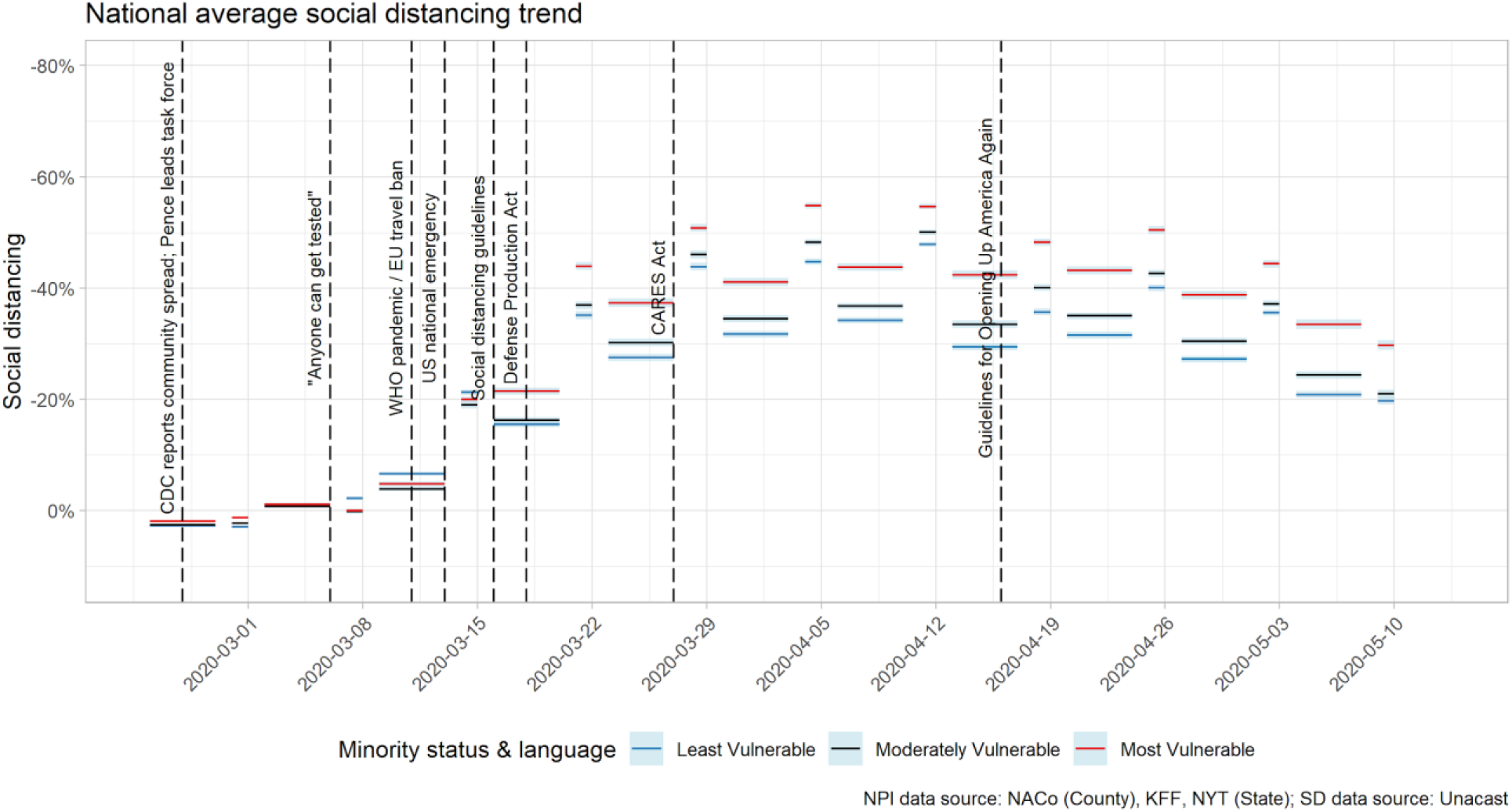
National average social distancing by CCVI minority status & language score.

**Table 6.** National Changes in Mobility (Social Distancing) across Three Time Periods for Average Weekdays and Weekends, by CCVI Theme Score. *Negative numbers represent a decrease in mobility, i.e. more social distancing than before; positive numbers represent an increase in mobility, i.e. less social distancing than before*

| Theme** | WHO Pandemic Declaration |  |  |  |  |  | OUAA Guidelines |  |  |  |  |  | State Reopening |  |  |  |  |  |
| --- | --- | --- | --- | --- | --- | --- | --- | --- | --- | --- | --- | --- | --- | --- | --- | --- | --- | --- |
|  | Weekday |  |  | Weekend |  |  | Weekday |  |  | Weekend |  |  | Weekday |  |  | Weekend |  |  |
|  | Least | Mod | Most | Least | Mod | Most | Least | Mod | Most | Least | Mod | Most | Least | Mod | Most | Least | Mod | Most |
|  | %Δ | %Δ | %Δ | %Δ | %Δ | %Δ | %Δ | %Δ | %Δ | %Δ | %Δ | %Δ | %Δ | %Δ | %Δ | %Δ | %Δ | %Δ |
| SES | -22.6* | -17.2* | -10.1* | -43.6* | -41.8* | -32.0* | 0.9* | 1.0* | 2.3* | 4.5* | 5.2* | 8.7* | 8.8* | 10.5* | 12.1* | 20.2* | 21.1* | 22.3* |
| HC & DISABILITY | -21.6* | -12.4* | -9.5* | -44.8* | -34.2* | -29.9* | 0.6* | 2.0* | 2.6* | 4.0* | 7.7* | 10.5* | 9.2* | 11.6* | 12.4* | 20.5* | 21.8* | 21.9* |
| MS & L | -14.3* | -15.5* | -20.4* | -32.9* | -37.1* | -44.0* | 2.7* | 1.8* | 0.6* | 7.8* | 7.4* | 4.2* | 10.7* | 10.6* | 9.6* | 20.3* | 21.6* | 20.7* |
| HT & T | -20.0* | -17.5* | -15.8* | -43.3* | -40.1* | -35.7* | 0.9* | 1.2* | 1.6* | 5.1* | 5.1* | 6.8* | 9.2* | 10.8* | 11.2* | 20.6* | 21.7* | 19.8* |
| EPI | -19.8* | -17.9* | -17.8* | -42.2* | -41.2* | -40.2* | 1.2* | 1.5* | 0.6* | 5.1* | 5.9* | 5.1* | 10.2* | 10.1* | 9.5* | 20.9* | 20.5* | 21.2* |
| HEALTH SYS | -19.3* | -19.1* | -16.6* | -40.9* | -42.7* | -39.5* | -0.3* | 1.5* | 2.7* | 4.7* | 5.1* | 6.9* | 9.3* | 10.4* | 10.3* | 20.4* | 21.3* | 20.8* |

\*\*SES: Socioeconomic Status, HC & DISABILITY: Household Composition & Disability, MS & L: Minority Status & Language, HT & T: Housing Type &
Transport, EPI: Epidemiological Factors, Health Sys: Health-System Factors
\*Significant at $p < 0.05$ ; otherwise <sup>(ns)</sup>, not significant

**Table 7.**
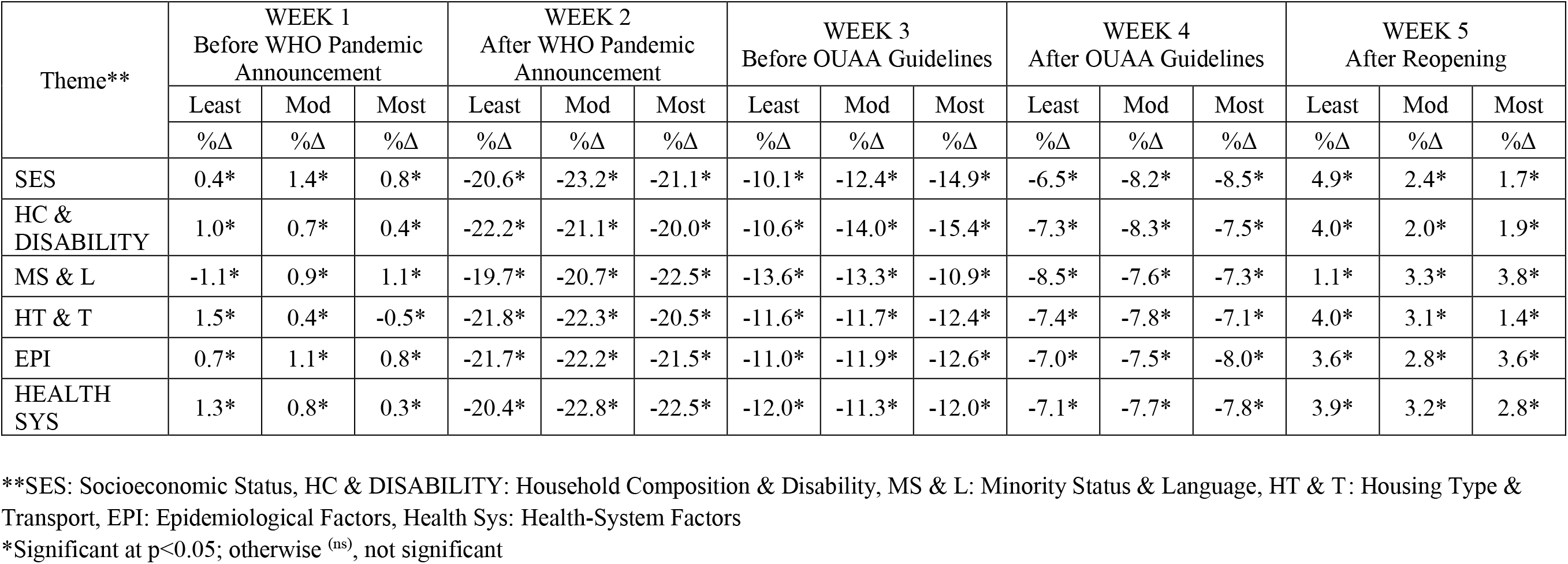
Difference in Level of Social Distancing between Average Weekend and Weekdays across Five Time Periods, by CCVI Theme Score.

| Theme** | WEEK 1<br>Before WHO Pandemic<br>Announcement |  |  | WEEK 2<br>After WHO Pandemic<br>Announcement |  |  | WEEK 3<br>Before OUAA Guidelines |  |  | WEEK 4<br>After OUAA Guidelines |  |  | WEEK 5<br>After Reopening |  |  |
| --- | --- | --- | --- | --- | --- | --- | --- | --- | --- | --- | --- | --- | --- | --- | --- |
|  | Least | Mod | Most | Least | Mod | Most | Least | Mod | Most | Least | Mod | Most | Least | Mod | Most |
|  | %Δ | %Δ | %Δ | %Δ | %Δ | %Δ | %Δ | %Δ | %Δ | %Δ | %Δ | %Δ | %Δ | %Δ | %Δ |
| SES | 0.4* | 1.4* | 0.8* | -20.6* | -23.2* | -21.1* | -10.1* | -12.4* | -14.9* | -6.5* | -8.2* | -8.5* | 4.9* | 2.4* | 1.7* |
| HC & DISABILITY | 1.0* | 0.7* | 0.4* | -22.2* | -21.1* | -20.0* | -10.6* | -14.0* | -15.4* | -7.3* | -8.3* | -7.5* | 4.0* | 2.0* | 1.9* |
| MS & L | -1.1* | 0.9* | 1.1* | -19.7* | -20.7* | -22.5* | -13.6* | -13.3* | -10.9* | -8.5* | -7.6* | -7.3* | 1.1* | 3.3* | 3.8* |
| HT & T | 1.5* | 0.4* | -0.5* | -21.8* | -22.3* | -20.5* | -11.6* | -11.7* | -12.4* | -7.4* | -7.8* | -7.1* | 4.0* | 3.1* | 1.4* |
| EPI | 0.7* | 1.1* | 0.8* | -21.7* | -22.2* | -21.5* | -11.0* | -11.9* | -12.6* | -7.0* | -7.5* | -8.0* | 3.6* | 2.8* | 3.6* |
| HEALTH SYS | 1.3* | 0.8* | 0.3* | -20.4* | -22.8* | -22.5* | -12.0* | -11.3* | -12.0* | -7.1* | -7.7* | -7.8* | 3.9* | 3.2* | 2.8* |

\*\*SES: Socioeconomic Status, HC & DISABILITY: Household Composition & Disability, MS & L: Minority Status & Language, HT & T: Housing Type & Transport, EPI: Epidemiological Factors, Health Sys: Health-System Factors
\*Significant at $p < 0.05$ ; otherwise <sup>(ns)</sup>, not significant

### Black Americans social distanced more on average than white Americans

Using social distancing data weighted by county population of race, we estimated that the average Black individual in the US social distanced significantly more than the average white individual at all time periods studied (Figure 4). The maximum level of social distancing reached on April 12th was greater for black Americans (58.8% versus 56%). For both black and white Americans, the change in weekday and weekend average social distancing matched national trends, with increased social distancing following the WHO announcement and decreased social distancing after the OUAA announcement and state reopenings (Table 8). The interaction effects of race on the change in social distancing before and after all three key events were significant (F(1, 6098) = 27.6, p<0.01); F(1, 6098) = 25.5, p<0.01); F(1, 6098) = 4.0, p<0.01)). The differences in weekday and weekend social distancing trends were similar, with little difference in the magnitude between black and white individuals over time (Table 9). The interaction effects of race on the difference in weekday and weekend social distancing were not significant for the weeks before and after the WHO announcement (week 1, week 2) and after states reopened (week 5), but were significant for the weeks before and after the OUAA announcement (week 3, week 4) (F(1, 6098) = 9.7, p<0.01).

**Figure 4.**
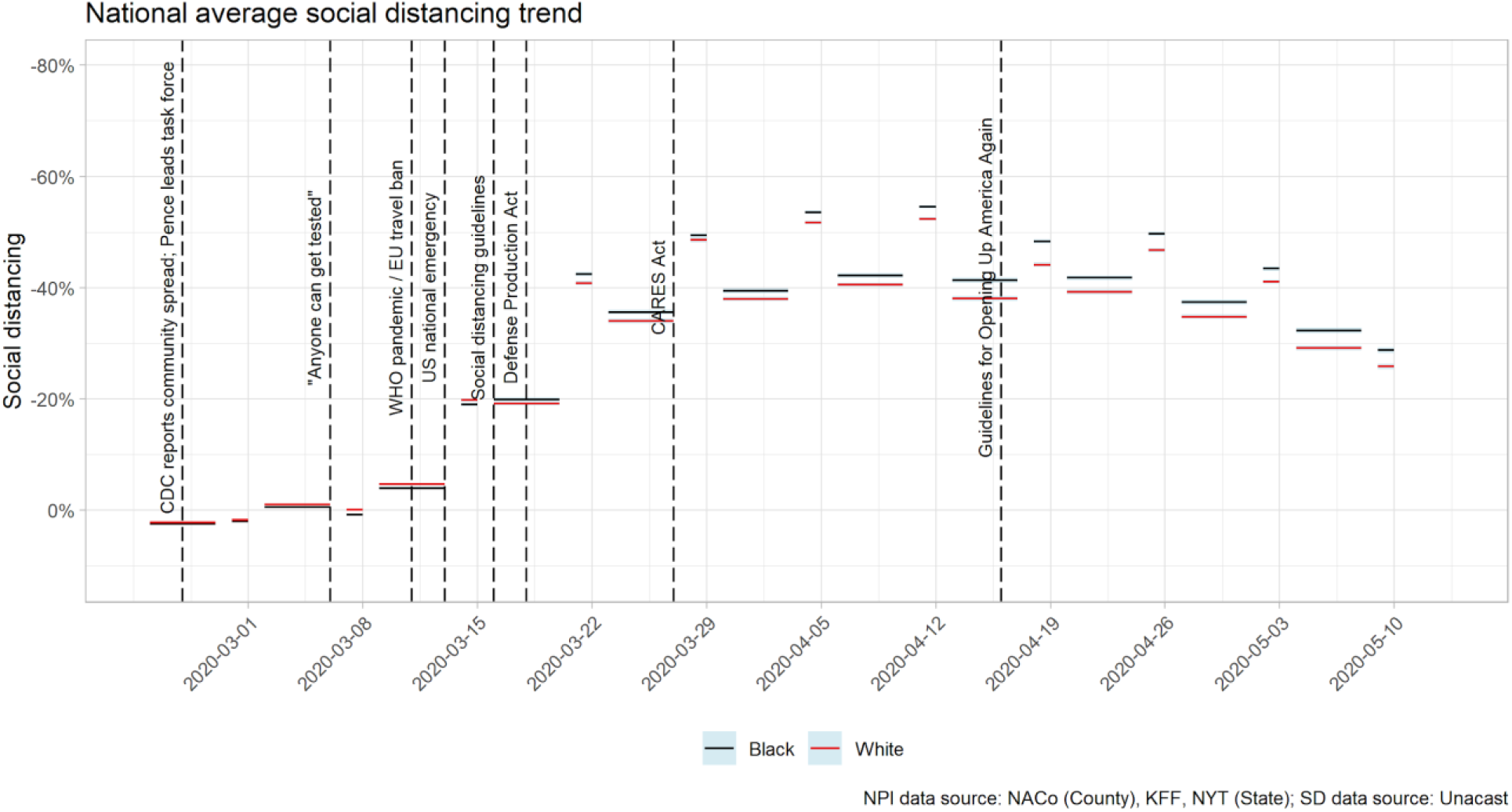
National Average Social Distancing by Race.

**Table 8.**
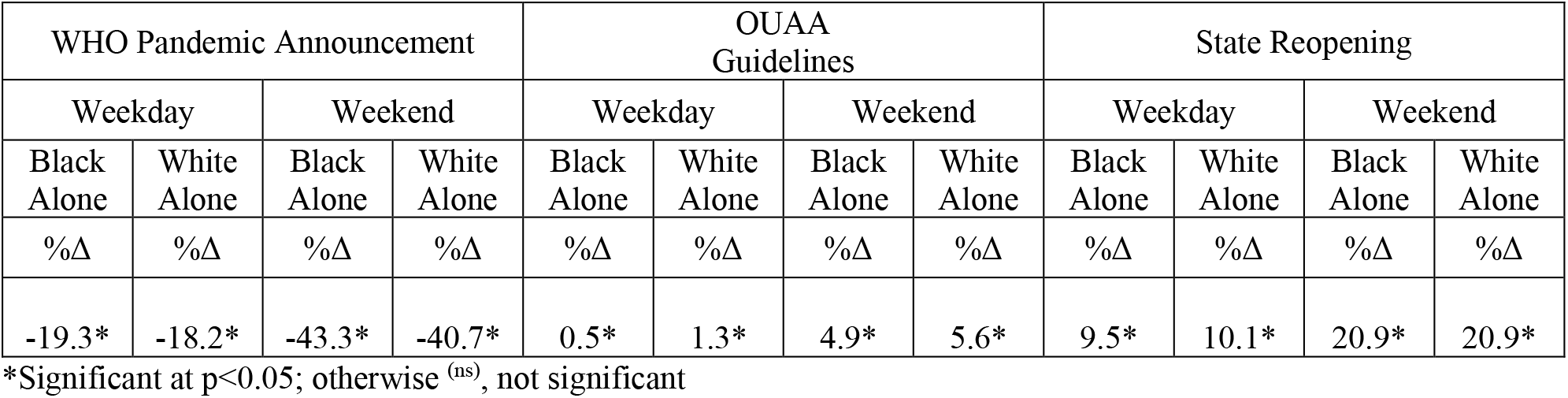
National Changes in Mobility (Social Distancing) across Three Time Periods for Average Weekdays and Weekends, by Race. *Negative numbers represent a decrease in mobility, i.e. more social distancing than before; positive numbers represent an increase in mobility, i.e. less social distancing than before*.

**Table 9.** Difference in Level of Social Distancing between Average Weekend and Weekdays across Five Time Periods, by Race. *Negative numbers represent less mobility, i.e., more social distancing, on weekends than weekdays; positive numbers represent more mobility, i.e., less social distancing, on weekends than weekdays*.

| WEEK 1<br>Before WHO<br>Pandemic<br>Announcement |  | WEEK 2<br>After WHO<br>Pandemic<br>Announcement |  | WEEK 3<br>Before OUAA<br>Guidelines |  | WEEK 4<br>After OUAA<br>Guidelines |  | WEEK 5<br>After Reopening |  |
| --- | --- | --- | --- | --- | --- | --- | --- | --- | --- |
| Black Alone | White Alone | Black Alone | White Alone | Black Alone | White Alone | Black Alone | White Alone | Black Alone | White Alone |
| %Δ | %Δ | %Δ | %Δ | %Δ | %Δ | %Δ | %Δ | %Δ | %Δ |
| 1.4* | 0.9* | -22.6* | -21.6* | -12.4* | -11.8* | -7.9* | -7.5* | 3.6* | 3.3* |

\*Significant at $p < 0.05$ ; otherwise <sup>(ns)</sup>, not significant

### 2016 Trump voters social distance on average less than Clinton voters

The average Trump voter (based on 2016 presidential election voting) social distanced significantly less than the average Clinton voter at all time periods studied (Figure 5). On April 12th, the maximum social distance achieved was greater for Clinton voters (58.6%) than for Trump voters (55.0%). Although overall social distancing rates differed by voting behavior, the change in weekday and weekend average social distancing again matched national trends for both groups, with increased social distancing following the WHO announcement and decreased social distancing after the OUAA announcement and state reopenings (Table 10). The interaction effects of 2016 presidential voting on the change in social distancing before and after all three key events were significant (F(1, 6072) = 228.5, p<0.01), (F(1, 6072) = 274.8, p<0.01), (F(1, 6072) = 61.9, p<0.01). Clinton voters on average increased social distancing by a greater magnitude following the WHO announcement, and decreased by a smaller magnitude following the OUAA guidelines and state reopenings. For both voter groups, social distancing matched national trends, with less social distancing on weekends early on and following state reopening, and more social distancing on the weekends in between (Table 11). The interaction effects of voter group on the difference in weekday and weekend social distancing were significant for the weeks before and after the WHO announcement (week 1, week 2) (F(1, 6072) = 22.7, p<0.01) and OUAA announcement (week 3, week 4) (F(1, 6072) = 38.0, p<0.01), however, were not significant after states reopened (week 5).

**Figure 5.**
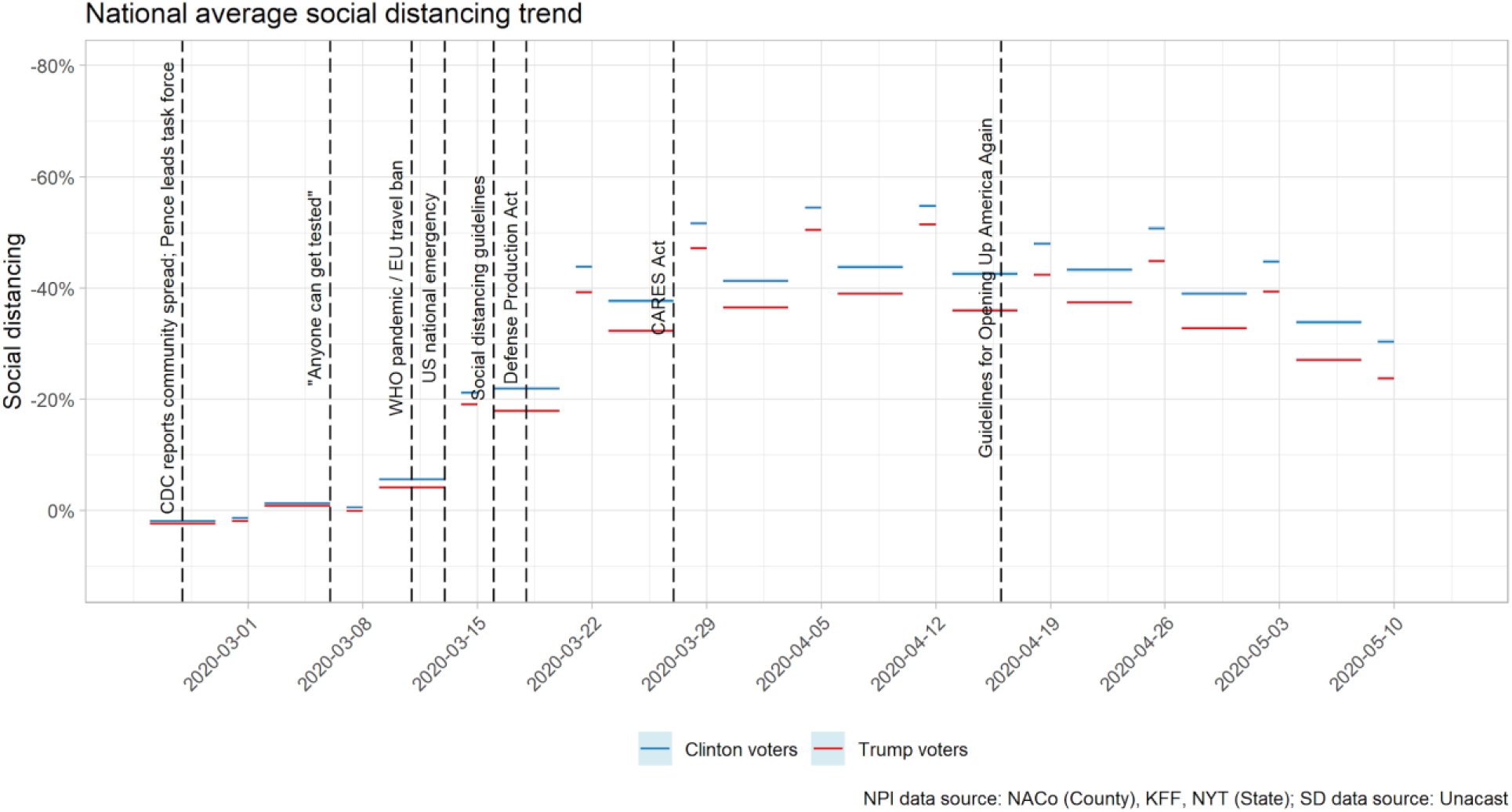
National Average Social Distancing by 2016 Presidential Election voters.

**Table 10.** National and Selected State-Level Changes in Mobility (Social Distancing) across Three Time Periods for Average Weekdays and Weekends, by 2016 Presidential Voting Behavior. *Negative numbers represent a decrease in mobility, i.e. more social distancing than before; positive numbers represent an increase in mobility, i.e. less social distancing than before*.

| WHO Pandemic Announcement |  |  |  | OUAA Guidelines |  |  |  | State Reopening |  |  |  |
| --- | --- | --- | --- | --- | --- | --- | --- | --- | --- | --- | --- |
| Weekday |  | Weekend |  | Weekday |  | Weekend |  | Weekday |  | Weekend |  |
| Dem | Rep | Dem | Rep | Dem | Rep | Dem | Rep | Dem | Rep | Dem | Rep |
| %Δ | %Δ | %Δ | %Δ | %Δ | %Δ | %Δ | %Δ | %Δ | %Δ | %Δ | %Δ |
| -20.6* | -17.1* | -43.2* | -39.4* | 0.5* | 1.6* | 4.0* | 6.6* | 9.4* | 10.4* | 20.4* | 21.1* |

\*Significant at $p < 0.05$ ; otherwise <sup>(ns)</sup>, not significant

**Table 11.** Difference in Level of Social Distancing between Average Weekend and Weekdays across Five Time Periods, by 2016 Presidential Votes. *Negative numbers represent less mobility, i.e., more social distancing, on weekends than weekdays; positive numbers represent more mobility, i.e., less social distancing, on weekends than weekdays*.

| WEEK 1<br>Before WHO<br>Pandemic<br>Announcement |  | WEEK 2<br>After WHO<br>Pandemic<br>Announcement |  | WEEK 3<br>Before OUAA<br>Guidelines |  | WEEK 4<br>After OUAA<br>Guidelines |  | WEEK 5<br>After Reopening |  |
| --- | --- | --- | --- | --- | --- | --- | --- | --- | --- |
| Dem | Rep | Dem | Rep | Dem | Rep | Dem | Rep | Dem | Rep |
| %Δ | %Δ | %Δ | %Δ | %Δ | %Δ | %Δ | %Δ | %Δ | %Δ |
| 0.7* | 1.0* | -21.9* | -21.3* | -11.0* | -12.4* | -7.5* | -7.4* | 3.4* | 3.3* |

\*Significant at $p < 0.05$ ; otherwise <sup>(ns)</sup>, not significant

### Manual employment sectors social distanced less

We computed the amount of social distancing by each employment sector. For clarity we pooled the social distance average for each sector and clustered based on social distancing level (Table 12).

**Table 12.** Employment Sector Clustering Results.

| Employment Sector | Found cluster based on k-means clustering |
| --- | --- |
| Arts Entertainment and Recreation | Social Distancing Most |
| Educational Services |  |
| Finance and Insurance |  |
| Information |  |
| Management of Companies and Enterprises |  |
| Professional Scientific and Technical Services |  |
| Real Estate and Rental and Leasing |  |
| Accommodation and Food Services | Mid social distancing |
| Administrative and Support and Waste Management and Remediation Services |  |
| Construction |  |
| Health Care and Social Assistance |  |
| Manufacturing |  |
| Other Services (except Public Administration) |  |
| Retail Trade |  |
| Transportation and Warehousing |  |
| Utilities |  |
| Wholesale Trade |  |
| Agriculture Forestry Fishing and Hunting | Low social distancing |
| Mining Quarrying and Oil and Gas Extraction |  |

We used k-means clustering to analyze differences in how various employment sectors social distanced on each day, and tallied the frequency. All sectors consistently (>60 days out of 77 available) classified into the same clusters, suggesting a systematic difference in the mobility patterns for employees in different sectors (Figure 6, Table 13). In particular, employees in sectors that closed early in the pandemic (Education services), or sectors that we can reasonably assume can work remotely (Information, Real Estate and Rental and Leasing, Professional Scientific and Technical Services, Management of Companies and Enterprises, Finance and Insurance) or have a high degree of freelance work (Arts, Entertainment and Recreation) clustered as social distancing the most. Sectors that often require on-site physical presence (Construction, Manufacturing, Utilities, Transportation and Warehousing, Administrative and Support, and Waste Management and Remediation Services) or customer-facing work (Retail Trades, Wholesale Trades, Accommodation and Food Services, Health Care and Social Assistance) social distanced less. Two employment sectors requiring on-site manual labor (Agriculture, Forestry, Fishing, and Hunting; and Mining, and Quarrying, and Oil and Gas Extraction) social distanced the least.

**Figure 6.**
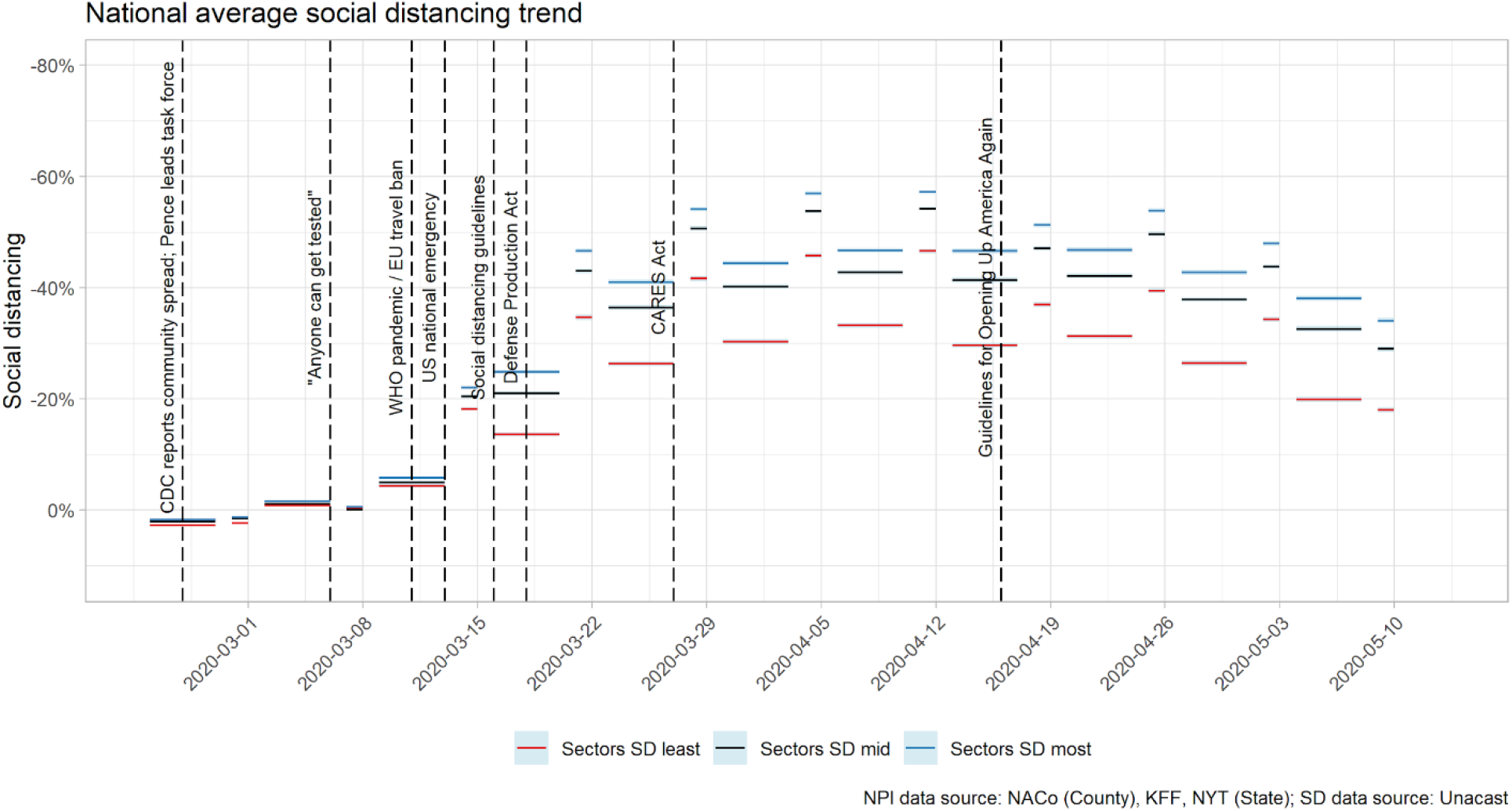
National Average Social Distancing by Employment Sector.

The interaction effects of employment sector clusters on the change in social distancing before and after all three key events were significant (F(2,8560)=138.1, p<0.01), (F(2,8560)=61.2, p<0.01), (F(2,8560)=42.2, p<0.01) (Table 14). The interaction effects of employment sector clusters on the difference in social distancing between weekdays and weekends for all three key events were not significant for WHO pandemic declaration (F(2,8560)=1.52, p=0.219), but were significant for the two later events (F(2,8560)=30.8, p<0.01), (F(2,8560)=15.7, p<0.01) (Table 15).

**Table 13.** Average Weekday and Weekend Level of Social Distancing across Five Time Periods, by Industry Sector.

| Sector | WEEK 1 |  | WEEK 2 |  | WEEK 3 |  | WEEK 4 |  | WEEK 5 |  |
| --- | --- | --- | --- | --- | --- | --- | --- | --- | --- | --- |
|  | Before WHO Pandemic Announcement |  | After WHO Pandemic Announcement |  | Before OUAA Guidelines |  | After OUAA Guidelines |  | After Reopening |  |
|  | Weekday | Weekend | Weekday | Weekend | Weekday | Weekend | Weekday | Weekend | Weekday | Weekend |
| Accommodation and Food Services | -1.22 (-1.29,-1.15)% | 0.16 (-0.01,0.32)% | -20.88 (-21.11,-20.65)% | -43.15 (-43.47,-42.83)% | -43.26 (-43.45,-43.07)% | -54.84 (-55.09,-54.58)% | -42.55 (-42.76,-42.34)% | -50.05 (-50.37,-49.74)% | -32.81 (-33.04,-32.57)% | -29.49 (-29.87,-29.12)% |
| Administrative and Support and Waste Management and Remediation Services | -1.18 (-1.24,-1.11)% | 0.11 (-0.03,0.26)% | -22.19 (-22.41,-21.98)% | -44.34 (-44.62,-44.05)% | -44.23 (-44.41,-44.06)% | -55.45 (-55.69,-55.21)% | -43.96 (-44.15,-43.77)% | -51.44 (-51.72,-51.16)% | -34.80 (-35.01,-34.58)% | -30.86 (-31.21,-30.51)% |
| Agriculture Forestry Fishing and Hunting | -1.19 (-1.28,-1.11)% | -1.47 (-1.63,-1.30)% | -13.78 (-13.99,-13.57)% | -35.39 (-35.76,-35.02)% | -33.37 (-33.56,-33.18)% | -46.45 (-46.75,-46.16)% | -30.85 (-31.07,-30.64)% | -39.82 (-40.14,-39.50)% | -18.84 (-19.06,-18.61)% | -17.73 (-18.09,-17.37)% |
| Arts Entertainment and Recreation | -1.59 (-1.66,-1.52)% | 0.09 (-0.09,0.26)% | -23.67 (-23.89,-23.44)% | -46.12 (-46.43,-45.82)% | -46.39 (-46.57,-46.21)% | -57.05 (-57.31,-56.79)% | -45.97 (-46.16,-45.77)% | -53.17 (-53.47,-52.88)% | -36.70 (-36.92,-36.47)% | -33.16 (-33.53,-32.79)% |
| Construction | -1.23 (-1.30,-1.16)% | -0.08 (-0.24,0.08)% | -20.61 (-20.83,-20.40)% | -42.35 (-42.65,-42.05)% | -42.38 (-42.56,-42.20)% | -53.67 (-53.91,-53.43)% | -41.46 (-41.66,-41.26)% | -48.64 (-48.94,-48.35)% | -31.71 (-31.92,-31.49)% | -27.70 (-28.04,-27.35)% |
| Educational Services | -1.45 (-1.52,-1.38)% | -0.70 (-0.86,-0.53)% | -25.17 (-25.41,-24.93)% | -47.08 (-47.39,-46.77)% | -47.09 (-47.29,-46.90)% | -57.26 (-57.53,-56.99)% | -47.14 (-47.35,-46.92)% | -54.47 (-54.79,-54.15)% | -38.65 (-38.90,-38.41)% | -35.23 (-35.63,-34.83)% |
| Finance and Insurance | -1.26 (-1.33,-1.19)% | -0.15 (-0.29,-0.00)% | -23.64 (-23.86,-23.42)% | -45.36 (-45.66,-45.06)% | -45.19 (-45.38,-45.01)% | -56.44 (-56.69,-56.18)% | -45.32 (-45.52,-45.11)% | -52.47 (-52.77,-52.16)% | -36.56 (-36.79,-36.33)% | -32.72 (-33.10,-32.34)% |
| Health Care and Social Assistance | -1.19 (-1.26,-1.12)% | -0.45 (-0.60,-0.30)% | -21.85 (-22.06,-21.64)% | -43.85 (-44.13,-43.56)% | -43.59 (-43.77,-43.42)% | -54.69 (-54.93,-54.45)% | -43.13 (-43.32,-42.93)% | -50.57 (-50.86,-50.28)% | -33.69 (-33.91,-33.47)% | -30.15 (-30.49,-29.80)% |
| Information | -2.44 (-2.51,-2.37)% | -1.66 (-1.83,-1.49)% | -27.31 (-27.54,-27.08)% | -48.74 (-49.03,-48.44)% | -48.83 (-49.01,-48.65)% | -58.61 (-58.86,-58.36)% | -49.29 (-49.49,-49.09)% | -56.02 (-56.32,-55.72)% | -40.75 (-40.97,-40.52)% | -36.20 (-36.57,-35.82)% |
| Management of Companies and Enterprises | -1.48 (-1.55,-1.40)% | -0.59 (-0.74,-0.44)% | -24.55 (-24.78,-24.32)% | -46.01 (-46.31,-45.71)% | -45.85 (-46.04,-45.67)% | -56.53 (-56.78,-56.27)% | -46.17 (-46.37,-45.97)% | -53.42 (-53.71,-53.13)% | -37.71 (-37.93,-37.48)% | -33.36 (-33.72,-33.00)% |
| Manufacturing | -0.98 (-1.05,-0.92)% | -0.31 (-0.45,-0.17)% | -19.93 (-20.13,-19.72)% | -41.66 (-41.94,-41.38)% | -40.62 (-40.79,-40.44)% | -52.74 (-52.97,-52.52)% | -39.83 (-40.03,-39.64)% | -47.71 (-47.98,-47.43)% | -30.21 (-30.42,-30.01)% | -26.69 (-27.00,-26.38)% |
| Mining Quarrying and Oil and Gas Extraction | -0.01 (-0.11,0.10)% | 2.01 (1.80,2.21)% | -13.16 (-13.40,-12.92)% | -33.05 (-33.49,-32.61)% | -32.87 (-33.08,-32.66)% | -46.82 (-47.15,-46.49)% | -32.03 (-32.25,-31.80)% | -38.61 (-38.97,-38.26)% | -22.05 (-22.28,-21.82)% | -18.57 (-18.96,-18.18)% |
| Other Services (except Public Administration) | -1.32 (-1.39,-1.25)% | -0.49 (-0.64,-0.33)% | -22.57 (-22.79,-22.35)% | -44.39 (-44.70,-44.09)% | -44.37 (-44.55,-44.18)% | -55.33 (-55.59,-55.08)% | -43.86 (-44.07,-43.66)% | -51.30 (-51.61,-51.00)% | -34.67 (-34.90,-34.44)% | -31.11 (-31.47,-30.74)% |
| Professional Scientific and Technical Services | -1.65 (-1.72,-1.58)% | -0.67 (-0.83,-0.52)% | -25.55 (-25.77,-25.33)% | -47.09 (-47.38,-46.80)% | -47.33 (-47.51,-47.15)% | -57.54 (-57.79,-57.30)% | -47.39 (-47.59,-47.19)% | -54.35 (-54.64,-54.06)% | -38.79 (-39.01,-38.56)% | -34.55 (-34.91,-34.18)% |
| Real Estate and Rental and Leasing | -1.50 (-1.57,-1.43)% | -0.09 (-0.25,0.08)% | -23.48 (-23.71,-23.26)% | -45.81 (-46.12,-45.51)% | -46.04 (-46.23,-45.85)% | -56.86 (-57.12,-56.61)% | -45.80 (-46.01,-45.60)% | -53.03 (-53.33,-52.72)% | -36.65 (-36.88,-36.41)% | -32.89 (-33.27,-32.51)% |
| Retail Trade | -1.15 (-1.22,-1.08)% | -0.12 (-0.27,0.04)% | -20.24 (-20.45,-20.02)% | -42.05 (-42.35,-41.74)% | -41.93 (-42.11,-41.75)% | -53.60 (-53.85,-53.36)% | -41.07 (-41.27,-40.87)% | -48.57 (-48.87,-48.27)% | -31.29 (-31.52,-31.07)% | -27.74 (-28.09,-27.39)% |
| Transportation and Warehousing | -0.90 (-0.97,-0.84)% | -0.01 (-0.15,0.14)% | -20.21 (-20.42,-20.01)% | -42.15 (-42.45,-41.85)% | -41.67 (-41.85,-41.48)% | -53.64 (-53.89,-53.39)% | -41.41 (-41.62,-41.21)% | -49.47 (-49.76,-49.17)% | -32.45 (-32.67,-32.23)% | -28.87 (-29.22,-28.53)% |
| Utilities | -1.00 (-1.07,-0.93)% | -0.02 (-0.19,0.15)% | -19.67 (-19.90,-19.44)% | -41.45 (-41.78,-41.12)% | -40.99 (-41.19,-40.80)% | -52.75 (-53.01,-52.48)% | -39.99 (-40.21,-39.77)% | -47.27 (-47.60,-46.93)% | -30.29 (-30.53,-30.05)% | -26.79 (-27.17,-26.41)% |
| Wholesale Trade | -1.14 (-1.20,-1.07)% | -0.19 (-0.33,-0.04)% | -21.52 (-21.73,-21.31)% | -43.48 (-43.77,-43.20)% | -42.79 (-42.97,-42.62)% | -54.43 (-54.67,-54.20)% | -42.32 (-42.52,-42.13)% | -49.94 (-50.22,-49.65)% | -33.22 (-33.43,-33.01)% | -29.34 (-29.67,-29.00)% |

**Table 14.** National Changes in Mobility (Social Distancing) across Three Time Periods for Average Weekdays and Weekends, by Industry Sector.

| Sector | WHO Pandemic Declaration |  | OUAA Guidelines |  | State Reopening |  |
| --- | --- | --- | --- | --- | --- | --- |
|  | Weekday | Weekend | Weekday | Weekend | Weekday | Weekend |
|  | %Δ | %Δ | %Δ | %Δ | %Δ | %Δ |
| Accommodation and Food Services | -19.66% | -43.31% | 0.71% | 4.78% | 9.74% | 20.56% |
| Administrative and Support and Waste Management and Remediation Services | -21.02% | -44.45% | 0.27% | 4.01% | 9.16% | 20.58% |
| Agriculture Forestry Fishing and Hunting | -12.59% | -33.92% | 2.52% | 6.63% | 12.02% | 22.09% |
| Arts Entertainment and Recreation | -22.08% | -46.21% | 0.43% | 3.87% | 9.27% | 20.01% |
| Construction | -19.38% | -42.27% | 0.92% | 5.02% | 9.76% | 20.95% |
| Educational Services | -23.72% | -46.38% | -0.04% | 2.79% | 8.49% | 19.24% |
| Finance and Insurance | -22.38% | -45.21% | -0.13% | 3.97% | 8.76% | 19.75% |
| Health Care and Social Assistance | -20.66% | -43.40% | 0.47% | 4.12% | 9.43% | 20.42% |
| Information | -24.87% | -47.08% | -0.46% | 2.59% | 8.54% | 19.82% |
| Management of Companies and Enterprises | -23.07% | -45.42% | -0.32% | 3.11% | 8.46% | 20.06% |
| Manufacturing | -18.94% | -41.35% | 0.78% | 5.04% | 9.62% | 21.02% |
| Mining Quarrying and Oil and Gas Extraction | -13.15% | -35.06% | 0.84% | 8.21% | 9.98% | 20.04% |
| Other Services (except Public Administration) | -21.25% | -43.91% | 0.50% | 4.03% | 9.19% | 20.19% |
| Professional Scientific and Technical Services | -23.90% | -46.42% | -0.06% | 3.19% | 8.60% | 19.80% |
| Real Estate and Rental and Leasing | -21.98% | -45.73% | 0.24% | 3.83% | 9.15% | 20.14% |
| Retail Trade | -19.09% | -41.93% | 0.86% | 5.04% | 9.78% | 20.83% |
| Transportation and Warehousing | -19.31% | -42.14% | 0.25% | 4.17% | 8.96% | 20.59% |
| Utilities | -18.67% | -41.43% | 1.01% | 5.48% | 9.70% | 20.48% |
| Wholesale Trade | -20.38% | -43.30% | 0.47% | 4.50% | 9.10% | 20.60% |

**Table 15.**
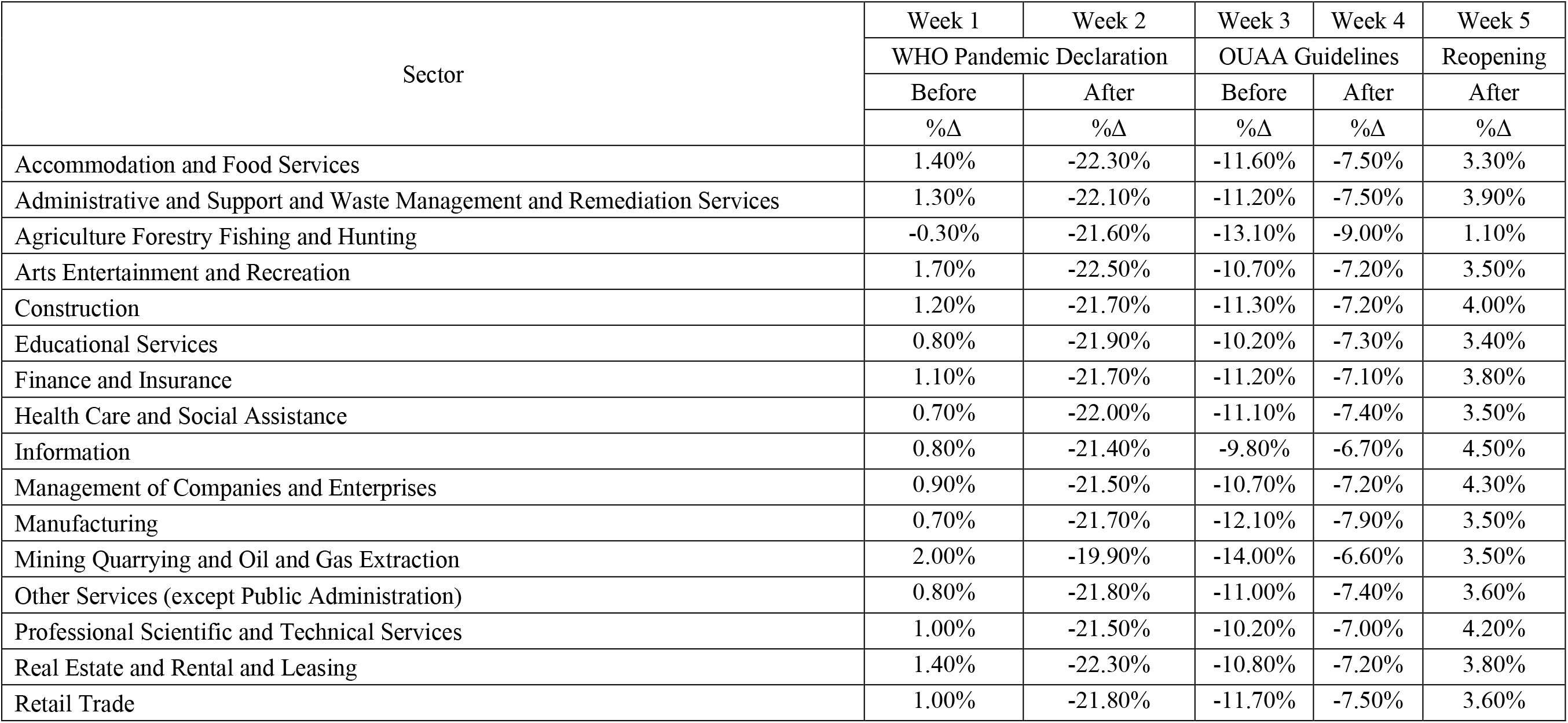
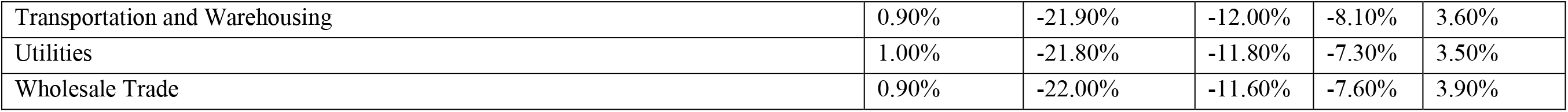
Difference in Level of Social Distancing between Average Weekend and Weekdays across Five Time Periods, by Industry Sector.

| Sector | Week 1 | Week 2 | Week 3 | Week 4 | Week 5 |
| --- | --- | --- | --- | --- | --- |
|  | WHO Pandemic Declaration |  | OUAA Guidelines |  | Reopening |
|  | Before | After | Before | After | After |
|  | %Δ | %Δ | %Δ | %Δ | %Δ |
| Accommodation and Food Services | 1.40% | -22.30% | -11.60% | -7.50% | 3.30% |
| Administrative and Support and Waste Management and Remediation Services | 1.30% | -22.10% | -11.20% | -7.50% | 3.90% |
| Agriculture Forestry Fishing and Hunting | -0.30% | -21.60% | -13.10% | -9.00% | 1.10% |
| Arts Entertainment and Recreation | 1.70% | -22.50% | -10.70% | -7.20% | 3.50% |
| Construction | 1.20% | -21.70% | -11.30% | -7.20% | 4.00% |
| Educational Services | 0.80% | -21.90% | -10.20% | -7.30% | 3.40% |
| Finance and Insurance | 1.10% | -21.70% | -11.20% | -7.10% | 3.80% |
| Health Care and Social Assistance | 0.70% | -22.00% | -11.10% | -7.40% | 3.50% |
| Information | 0.80% | -21.40% | -9.80% | -6.70% | 4.50% |
| Management of Companies and Enterprises | 0.90% | -21.50% | -10.70% | -7.20% | 4.30% |
| Manufacturing | 0.70% | -21.70% | -12.10% | -7.90% | 3.50% |
| Mining Quarrying and Oil and Gas Extraction | 2.00% | -19.90% | -14.00% | -6.60% | 3.50% |
| Other Services (except Public Administration) | 0.80% | -21.80% | -11.00% | -7.40% | 3.60% |
| Professional Scientific and Technical Services | 1.00% | -21.50% | -10.20% | -7.00% | 4.20% |
| Real Estate and Rental and Leasing | 1.40% | -22.30% | -10.80% | -7.20% | 3.80% |
| Retail Trade | 1.00% | -21.80% | -11.70% | -7.50% | 3.60% |
| Transportation and Warehousing | 0.90% | -21.90% | -12.00% | -8.10% | 3.60% |
| Utilities | 1.00% | -21.80% | -11.80% | -7.30% | 3.50% |
| Wholesale Trade | 0.90% | -22.00% | -11.60% | -7.60% | 3.90% |

## Discussion

Until a vaccine is widely available, social distancing in some form will continue to be one of the only effective approaches for preventing the spread of the coronavirus. Despite its importance, social distancing practice remains sensitive to shifts in announcements, policy enactments, and individual and community-level characteristics. Using mobility data as a proxy for social distancing, we found that social distancing in the US increased significantly throughout March and peaked on the Sunday (April 12) before the US government announced the OUAA guidelines. After the peak, despite continued growth in the total number of new cases, we observed significant declines in social distancing nationally – initially in the absence of guidance and policies permitting movement, but with additional significant declines following the OUAA announcement and the relaxation of individual state policies restricting movement. Interestingly, we found that shifts in social distancing began prior to specific policy enactments (whether to restrict or to relax movements), suggesting that people may shift behavior in anticipation of policy changes. We also observe that social distancing was much greater on weekends than weekdays throughout March, suggesting weekday activities (presumably employment-related) were still limiting how much people could social distance. By early May this gap disappeared and reversed, with even less social distancing on weekends than weekdays. This suggests a fundamental shift in how much caution individuals were choosing to exercise in their free time.

These findings have important implications for the future of the pandemic response. First, social distancing behaviors change over time, regardless of specific policy mandates. The observed decline in social distancing prior to any policies allowing increased movement could be due to individual social distancing fatigue leading to reduced precautions in behavior, or to public discussion of predicted reopening policies triggering preemptive behavior change. During outbreaks, protective behaviors can be influenced by news coverage on the current reported severity (Andersen, 2020; Fenichel, Kuminoff, & Chowell, 2013; Wong & Sam, 2010) or mismatched expectations between individual perception and actual policy mandates (i.e. policies lasted longer than expected) (Briscese, Lacetera, Macis, & Tonin, 2020).

While this analysis cannot show what caused the decline in social distancing, the fact that individuals change their behaviors before policies are enacted suggests the need to keep a close pulse on social distancing behaviors so that policy can appropriately respond to behavior – or, more importantly, accurately anticipate it, to enforce social distancing as needed. Research suggests that risk perception and community norms are key predictors of social distancing behavior (Charles et al., 2020). Mitigating the negative costs of social distancing (e.g., providing monetary compensation for lost income) (Bodas & Peleg, 2020; Wright et al., 2020) or targeting risk communication to those who may feel less impacted (e.g., wealthy individuals) can improve social distancing compliance (Bodas & Peleg, 2020). Responding to observed changes in behavior requires policy, messaging, and communication that will appeal to these behavioral drivers and consider expectations of social distancing compliance.

Second, policy signaling can influence behavior. Although social distancing declined prior to reopening decisions, there remains an observed relationship between national policy announcements and shifts in behavior. Social distancing did not significantly increase until the week following the declaration of the pandemic by the WHO and the national emergency declaration by the White House (and this preceded the release of social distancing guidelines by the CDC). Social distancing significantly declined nationally following the release of the OUAA guidelines and after the first state reopenings. This suggests that individuals will adjust their behavior in response to national policy announcements, even when these do not include specific mandates or relaxations of mandates. Before announcing changes to come, public officials must recognize that individuals may shift their behavior because of the mere announcement itself, even if the policy does not suggest doing so.

Third, the shifting difference between weekend and weekday social distancing hints at the potential influence of employment barriers to distancing, as well as changes in social distancing intent over time. That for the initial 1.5 months of the pandemic social distancing was higher on the weekends, when individuals have more leisure time, suggests that it was structural barriers, such as requirements to perform essential work, or lack of paid sick leave, that led to lower levels of social distancing on weekdays. In future waves of the pandemic, it will be important to build in more opportunities for weekday social distancing, such as expanding work from home opportunities and allowing for paid sick leave across sectors, as well as building out protective measures for those who must work. The recent shift to less weekend social distancing implies that in later months of the pandemic, individuals may be travelling more during their free time, adding additional evidence that intent to social distance is declining. This is a key signal for policymakers: while social distancing has declined overall as states have reopened, this appears not only a product of people returning to work, but of people choosing to be more active on their weekends.

Differences emerge in social distancing among subgroups of the US population. More vulnerable counties, as measured by the CCVI, social distance less than less vulnerable counties, particularly counties that are poorer, have more households with elderly, young, and disabled members, those with limited transport access and crowded living arrangements, and those with less healthcare-system resources. More vulnerable counties also tend to have a greater within-week difference in weekday vs. weekend social distancing, but after reopening, this difference shrinks. In contrast, after reopening, less vulnerable counties have a greater within-week disparity, suggesting they experienced a disproportionate decline in voluntary weekend social distancing. The four vulnerability themes that differ the most in rates of social distancing suggest that vulnerable counties encounter structural barriers to social distancing, especially during weekdays. These may include lower-paying jobs in essential services that do not allow work from home, and a need to travel greater distances to get to employment. Findings are similar to existing research indicating that poverty reduces social distancing compliance (Wright et al., 2020). Economic barriers to social distancing must be addressed to enable better social distancing in these communities. Recognizing that these barriers will likely persist, it is also important to prioritize other interventions to combat the spread of COVID-19 in these communities, such as ensuring safe work environments and adequate and accessible personal protective equipment.

While in general those that are more vulnerable social distance less, the exception to this trend is counties that are more vulnerable in minority status and language, who consistently social distance more than communities that are less vulnerable in this regard. We conducted additional analysis on the relationship between race and social distancing by weighting social distancing data directly with subpopulation estimates of county racial breakdown. We again found that Black Americans social distance more than white Americans. Taken together, the results suggest that there are influences that we have not accounted for that contribute to demographic differences in social distancing behavior, such as differences in urban/rural settings, preferred information sources, risk perception, and community social norms.

In addition to community (i.e., vulnerability) and demographic (i.e., race) differences that may influence social distancing, there also appear to be differences based on the beliefs people hold. Clinton voters social distance more on average than Trump voters, similar to findings of previous research (Allcott et al., 2020; Andersen, 2020; Painter & Qiu, 2020). Past research suggests Democrats have high perceptions of the risk and severity of COVID-19 and attach greater perceived importance to social distancing (Allcott et al., 2020). Partisan differences may also be driven by perceived credibility of politicians and messaging (Allcott et al., 2020; Andersen, 2020; Painter & Qiu, 2020). Combined with the findings on vulnerability, this ideological divide in social distancing behaviors suggests that there is a need for more targeted messaging around social distancing tailored to the specific barriers an individual faces. Messages should come from diverse messengers, as crisis communication shows the most effective messaging comes from sources an individual feels a connection with, which will vary by the population targeted (Heath, Lee, & Ni, 2009). While some Americans may face structural barriers that prevent them from social distancing, such as jobs in essential services, others may be doing because of beliefs such as lower risk perception about the value of social distancing or different perceived community norms (Allcott et al., 2020; Charles et al., 2020). For these individuals, messaging should focus on the value of social distancing in keeping them and their communities safe.

Lastly, we look at different employment sectors. Employees in sectors that typically require less customer-facing interaction, such as information and professional services, and sectors that are currently shut down, such as educational services, social distance more. By contrast, employees in sectors such as manufacturing, retail, and accommodation and food services social distance less throughout. The lowest social distancing group is those in agricultural sectors and mining. This aligns with research showing that social distancing’s impact on employment productivity also varies by industry and geographic location, requiring different levels of compensation to offset losses for continued business operation (Koren & Pető, 2020). In addition to laws focused on job security, income replacement, and business relief (Koren & Pető, 2020; Rothstein & Talbott, 2007), policymakers need to consider additional workplace protection measures such as masks with continued reopening, especially for sectors unable to maintain some level of social distancing. This is critical given that risk of transmission can also vary by industry; work-related transmission contributed substantially to the early growth in COVID-19 cases throughout Asia (Lan, Wei, Hsu, Christiani, & Kales, 2020).

This analysis has several limitations. The most apparent is that many vulnerability dimensions and population characteristics are confounded. Second, because policy effects are likely cumulative, it is difficult to tease out individual policy effects. The selected national announcements may also not have any causal relationship with the observed shifts in social distancing behavior and may not reflect the true first shift in increasing or decreasing social distancing behavior. However, observational data show a clear large difference in the magnitude of social distancing before and after these time periods, in line with broad changes in the national discourse, suggesting a certain relationship. Lastly, we have not attempted to fully capture the perceptual drivers and other potential signals such as news, misinformation campaigns, and social norms. The reported results are therefore only observational in nature and cannot quantify the true causal effects of policies on social distancing behavior. However, our findings show that different communities respond in social distancing differently. More research and analysis is needed to understand how different policy approaches to restrictions and relaxation impact progress along the epidemic curve, and to identify which drivers, such as policy type or trust in government, are to be found in the causal pathway. Research is also needed to explore the causal relationship between specific policies and behaviors as well as how the described trends vary at the state level and in response to state and local policy shifts.

The social distancing rates based on Unacast’s mobility measurement assume that the population studied uses smartphones with such apps activated. Therefore, it may not model a representative sample of the movement of all individuals in a county. However, the data are considered a reasonable proxy for social distancing given the high levels of smartphone penetration in the US, and that measures are calculated based on distance traveled rather than location. Social distancing measures modeled from location data also make behavioral assumptions that may not be universal (e.g., all individuals having one “home” location throughout the pandemic, rather than relocating to family homes or other locations) (Walle, 2020). The definition of prepandemic baseline was determined by Unacast without our input; however, we believe Unacast has made a reasonable choice.

Interpretation of analyses may be limited by the availability of samples and their granularity. For example, vulnerability is ranked per county relative to all other counties at a national level, resulting in a heterogeneous distribution of low, moderate, and highly vulnerable counties from the perspective of the country. Here we have presented analysis at the national level but similar analyses were also performed (not shown). In a single state, though, there may be very few or even no low-vulnerability counties. This results in greater uncertainty of how representative a social distance estimate may be for finer geographical levels. Both race and political-affiliation data were aggregated as a per-category population-weighted average of county social distancing behavior. This requires an assumption that individual social distancing behaviors in these groupings follow that of a normal distribution (and similar to the county average).

While future analysis is needed to identify the specific factors that cause individuals to shift their social distancing behavior, our observational analysis of social distancing changes over time suggests that that behavior may be sensitive to both policies themselves, anticipation of policies to come, as well as individual and community-level characteristics that can make an individual more or less likely to social distance. To curb the spread of the virus, these factors must be acknowledged and addressed in future policies.

## Data Availability

Data are available with their respective owners and repositories.

https://github.com/CSSEGISandData/COVID-19

https://precisionforcovid.org/ccvi

https://dataverse.harvard.edu/dataset.xhtml?persistentId=doi:10.7910/DVN/VOQCHQ

https://www.bls.gov/cew/about-data/home.htm

https://www.unacast.com/data-for-good

## Acknowledgement

We thank Unacast and its Data for Good initiative for providing the social distancing data for humanitarian efforts and global health causes.

---

**Appendix Table 1.** States reopening on or before May 10.

| Week | States Reopening |
| --- | --- |
| 4/20-4/26 | Alaska, Colorado, South Carolina, Vermont, Georgia, Hawaii, Oklahoma, Michigan, Minnesota, Montana |
| 4/27-5/3 | Alabama, Tennessee, Louisiana, Utah, Texas, Iowa, North Dakota, Idaho, West Virginia, Wisconsin, Maine, Ohio, Nevada, Wyoming, Illinois, Mississippi |
| 5/4-5/10 | North Carolina, Missouri, Florida, Kansas, Nebraska, Pennsylvania, Rhode Island, Indiana, Arizona, Washington, California, Delaware |

**Appendix Figure 1.**
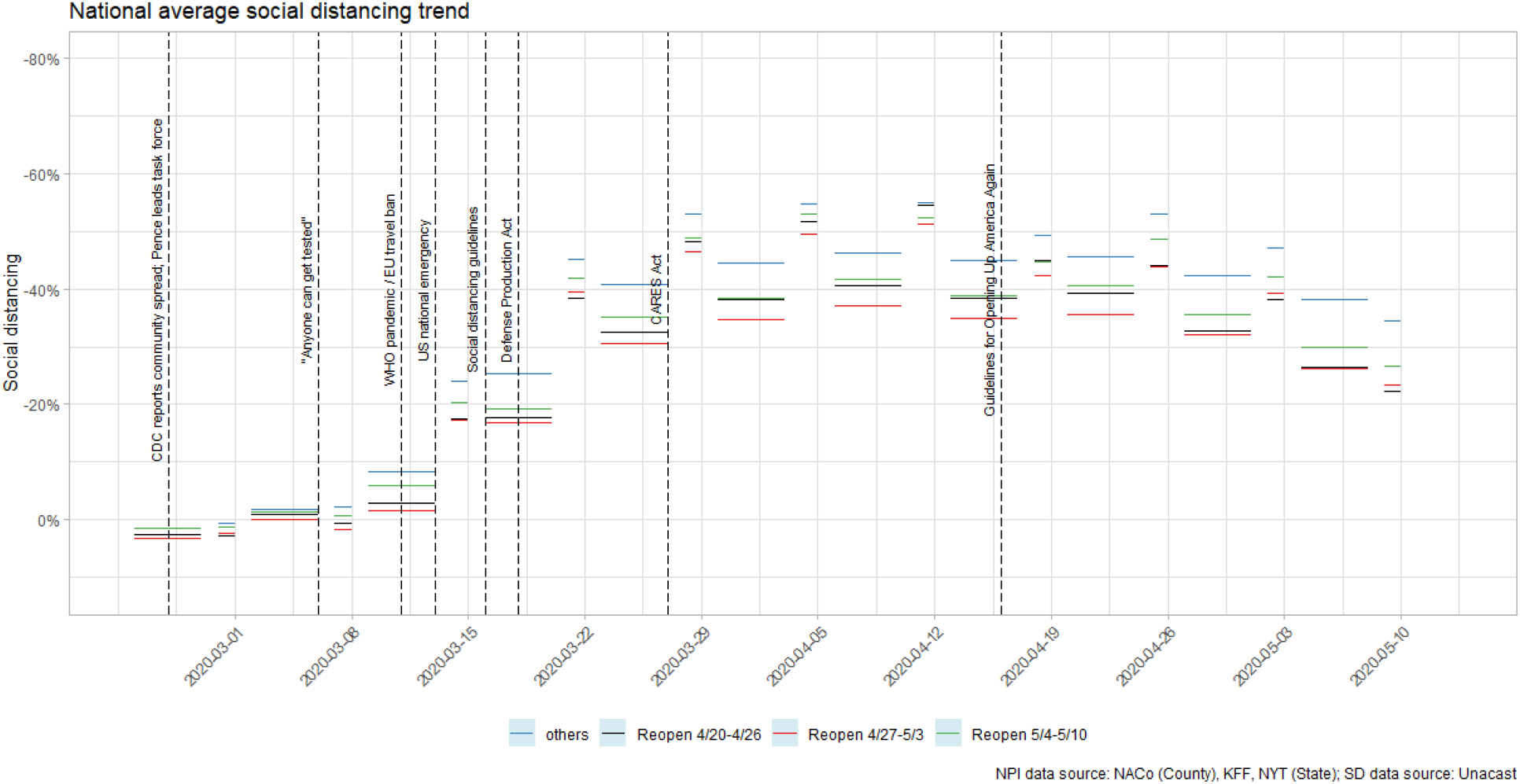
Social distancing by reopening groups on or before May 10^th^.

## Notes

### Competing Interest Statement

The authors have declared no competing interest.

### Funding Statement

No external funding was received.

### Author Declarations

All data sets were deidentified and aggregated before we received them. We received Unacast's deidentified social distancing data already aggregated at the level of counties, which cannot be traced to individuals. Unacast has an explicit privacy and consent policy (https://www.unacast.com/opt-out) stating that mobile phone users have opt-in consent for the collection of location data from mobile devices. Since the current study is a secondary analysis of existing, deidentified datasets obtained at the county-level, it did not require IRB approval. Surgo Foundation approved the study.

